# Preliminary Efficacy of the NVX-CoV2373 Covid-19 Vaccine Against the B.1.351 Variant

**DOI:** 10.1101/2021.02.25.21252477

**Authors:** Vivek Shinde, Sutika Bhikha, Zaheer Hoosain, Moherndran Archary, Qasim Bhorat, Lee Fairlie, Umesh Lalloo, Mduduzi S. L. Masilela, Dhayendre Moodley, Sherika Hanley, Leon Fouche, Cheryl Louw, Michele Tameris, Nishanta Singh, Ameena Goga, Keertan Dheda, Coert Grobbelaar, Gertruida Kruger, Nazira Carrim-Ganey, Vicky Baillie, Tulio de Oliveira, Anthonet Lombard Koen, Johan J. Lombaard, Rosie Mngqibisa, As’ad Ebrahim Bhorat, Gabriella Benadé, Natasha Lalloo, Annah Pitsi, Pieter-Louis Vollgraaff, Angelique Luabeya, Aliasgar Esmail, Friedrich G. Petrick, Aylin Oommen Jose, Sharne Foulkes, Khatija Ahmed, Asha Thombrayil, Lou Fries, Shane Cloney-Clark, Mingzhu Zhu, Chijioke Bennett, Gary Albert, Emmanuel Faust, Joyce S. Plested, Andreana Robertson, Susan Neal, Iksung Cho, Greg M. Glenn, Filip Dubovsky, Shabir A. Madhi, for the 2019nCoV-501 Study Group

**Affiliations:** Novavax, Inc., 20 Firstfield Road, Gaithersburg, MD, USA; South African Medical Research Council, Vaccines and Infectious Diseases Analytics Research Unit, Faculty of Health Sciences, University of the Witwatersrand, Johannesburg, South Africa; Josha Research Centre, Bloemfontein, Free State, South Africa; Paediatric Infectious Diseases Unit, University of KwaZulu-Natal, Durban, South Africa; Soweto Clinical Trials Centre, Johannesburg, South Africa; Wits Reproductive Health and HIV Institute, University of the Witwatersrand, Johannesburg, South Africa; Respiratory and Critical Care Unit, Nelson R. Mandela School of Medicine, University of KwaZulu-Natal, Durban, South Africa; Setshaba Research Centre, Tshwane, South Africa; Department of Obstetrics and Gynaecology, University of KwaZulu-Natal, Durban, South Africa; Centre Aids Prevention Research South Africa (CAPRISA), University of KwaZulu-Natal, Durban, South Africa; Limpopo Clinical Research Initiative, Rustenburg, North-West, South Africa; Madibeng Centre for Research, Department of Family Medicine, School of Health, University of Pretoria, Pretoria, South Africa; South African TB Vaccine Initiative, University of Cape Town, Cape Town, South Africa; Health Systems Research Unit and HIV Prevention Research Unit, South African Medical Research Council, Cape Town, South Africa; Centre for Lung Infection and Immunity, Division of Pulmonology, Department of Medicine and UCT Lung Institute, University of Cape Town, Cape Town, South Africa; Aurum Institute, University of Pretoria, Pretoria, South Africa; MERC Research, Middelburg, South Africa; PEERMED Clinical Trial Centre, Kempton Park, South Africa; Kwazulu-Natal Research Innovation and Sequencing Platform (KRISP), University of KwaZulu-Natal, Durban, South Africa

## Abstract

**Background:** The emergence of severe acute respiratory syndrome coronavirus 2 (SARS-CoV-2) variants threatens progress toward control of the Covid-19 pandemic. Evaluation of Covid-19 vaccine efficacy against SARS-CoV-2 variants is urgently needed to inform vaccine development and use.

**Methods:** In this phase 2a/b, multicenter, randomized, observer-blinded, placebo-controlled trial in South Africa, healthy human immunodeficiency virus (HIV)-negative adults (18 to 84 years) or medically stable people living with HIV (PLWH) (18 to 84 years) were randomized in a 1:1 ratio to receive two doses, administered 21 days apart, of either NVX-CoV2373 nanoparticle vaccine (5 µg recombinant spike protein with 50 µg Matrix-M1 adjuvant) or placebo. The primary endpoints were safety and vaccine efficacy ≥7 days following the second dose against laboratory-confirmed symptomatic Covid-19 in previously SARS-CoV-2 uninfected participants.

**Results:** A total of 4387 participants were randomized and dosed at least once, 2199 with NVX-CoV2373 and 2188 with placebo. Approximately 30% of participants were seropositive at baseline. Among 2684 baseline seronegative participants (94% HIV-negative; 6% PLWH), there were 15 and 29 predominantly mild to moderate Covid-19 cases in NVX-CoV2373 and placebo recipients, respectively; vaccine efficacy was 49.4% (95% confidence interval [CI]: 6.1 to 72.8). Efficacy in HIV-negative participants was 60.1% (95% CI: 19.9 to 80.1), and did not differ by baseline serostatus. Of the primary endpoint cases with available whole genome sequencing, 38 (92.7%) of 41 were the B.1.351 variant. Post-hoc vaccine efficacy against B.1.351 was 51.0% (95% CI: - 0.6 to 76.2) in HIV-negative participants. Among placebo recipients, the incidence of symptomatic Covid-19 was similar in baseline seronegative vs baseline seropositive participants during the first 2 months of follow-up (5.3% vs 5.2%). Preliminary local and systemic reactogenicity were primarily mild to moderate and transient, and higher with NVX-CoV2373; serious adverse events were rare in both groups.

**Conclusions:** The NVX-CoV2373 vaccine was efficacious in preventing Covid-19, which was predominantly mild to moderate and due to the B.1.351 variant, while evidence of prior infection with the presumptive original SARS-CoV-2 did not confer protection against probable B.1.351 disease. (Funded by Novavax, The Bill and Melinda Gates Foundation, and the Coalition for Epidemic Preparedness Innovations; ClinicalTrials.gov number, NCT04533399)

## INTRODUCTION

The coronavirus disease 2019 (Covid-19) pandemic, caused by the emergence of a novel severe acute respiratory syndrome coronavirus 2 (SARS-CoV-2), has resulted in over 109 million documented cases and 2.4 million deaths worldwide as of February 17, 2021.^1,2^ Vaccination remains a cornerstone of control strategies. Current vaccines primarily target the Covid-19 spike protein based on the prototype Wuhan strain.^3^ The mRNA vaccines (BNT162b2 and mRNA-1273) have demonstrated vaccine efficacy of 94% to 95%^4,5^, and vector-based vaccines reported vaccine efficacy of 71% (pooled) for ChAdOx1-nCoV19, 92% for Gam-COVID-Vac, and 66% for Ad26.COV2.S.^6,7^

We report on a recombinant, *Spodoptera frugiperda* (Sf9) insect cell/baculovirus system derived, SARS-CoV-2 nanoparticle vaccine (NVX-CoV2373) comprised of full-length, pre-fusion trimers of spike glycoprotein (prototype Wuhan sequence), co-formulated with a saponin-based adjuvant, Matrix-M1™.^8,9^ In an ongoing randomized, placebo-controlled, phase 1/2 trial in healthy adults, NVX-CoV2373, administered in a two-dose regimen 21 days apart, had an acceptable safety profile; was associated with a strong, Th1-biased, antigen-specific polyfunctional CD4+ T-cell response; and induced neutralizing antibody responses 4-fold higher than levels in convalescent sera from predominantly moderate to severe Covid-19 cases.^10^

Recent reports from the United Kingdom (UK), Brazil, and South Africa on the emergence of the B.1.1.7, P1, and B.1.351 (N501Y.V2) variants, respectively, confirm the acquisition of mutations in key antigenic sites in the receptor binding domain (RBD) and N-terminal domain of the spike protein.^11-16^ These antigenic changes may render naturally acquired or vaccine-derived immunity to prototype-like virus less effective against subsequent infection with variant viruses.^12,16-18^ Here, we describe early findings on the primary efficacy endpoint and preliminary safety of a randomized, observer-blinded, placebo-controlled, phase 2a/b trial of NVX-CoV2373 in 4406 participants in South Africa during a period of predominant circulation of B.1.351 variant viruses.

## METHODS

### Trial Objectives, Participants and Oversight

In this randomized, observer-blinded, placebo-controlled phase 2a/b trial, we assessed the safety and efficacy of two doses of NVX-CoV2373, administered 21 days apart. Participants were healthy human immunodeficiency virus (HIV)-negative adults 18 to 84 years of age or a subgroup of medically stable people living with HIV (PLWH) 18 to 64 years of age. As a safety measure, enrollment was staggered into Stage 1 and Stage 2 for both HIV-negative and PLWH groups, with progression from Stage 1 to Stage 2 in each group requiring favorable review of safety data through Day 7 from the prior stage against prespecified vaccination pause rules (**Tables S4, S6, and S7**). Key exclusion criteria were chronic administration of immunosuppressive therapy, autoimmune or immunodeficiency disease (except for medically stable PLWH), history of prior or current symptomatic Covid-19, or nucleic acid amplification test (NAAT)-confirmed SARS-CoV-2 infection (hereafter “confirmed”) performed as part of screening within 5 days prior to anticipated initial dosing. All participants provided written informed consent prior to enrollment. Further details of the trial design, conduct, oversight, and analyses are provided in the protocol, statistical analysis plan, and Supplementary Appendix (available with the full text of this article at NEJM.org).

NVX-CoV2373 was developed by Novavax, which designed and sponsored the trial, with funding support from the Bill and Melinda Gates Foundation and the Coalition for Epidemic Preparedness Innovations. The trial protocol was approved by the South African Health Products Regulatory Authority (SAHPRA; Ref 20200420) and Institutional Ethics Review Boards and registered in Clinicaltrials.gov (NCT045333990 and the Pan African Clinical trials Registry (PACTR202009726132275). Safety oversight, including for specific vaccination pause rules, was performed by an independent safety monitoring committee.

Trial data were available to all authors, who confirmed the accuracy and completeness of the data and the fidelity of the trial to the protocol.

### Trial Procedures

Participants were randomly assigned in a 1:1 ratio to receive two intramuscular injections, 21 days apart, of either NVX-CoV2373 (5 µg recombinant spike protein with 50 µg Matrix-M1 adjuvant) or saline placebo injection volume, 0.5 mL), administered by unblinded staff not otherwise involved with other study procedures or data collection. All other study staff and trial participants remained blinded to treatment assignment.

Participants were scheduled for in-person follow-up visits on Days 7, 21, 35, and Months 3, 6, and 12 (phone call only) to collect vital signs, adverse events, concomitant medication changes, and blood for immunogenicity analyses.

### Safety Assessments

The primary safety endpoints were occurrence of unsolicited adverse events (medically attended, serious, and those of special interest [**Tables S1 and S2**]) through Day 35 and solicited local and systemic adverse events evaluated via reactogenicity diary for 7 days following each vaccination (**Tables S3 and S4**). Safety follow-up is ongoing through Month 12.

### Efficacy Assessments

The primary efficacy endpoint was confirmed symptomatic mild, moderate, or severe Covid-19 (hereafter “symptomatic Covid-19”) in participants seronegative to SARS-CoV-2 at baseline occurring 7 days after receipt of the second study vaccine (ie, after Day 28) (**Table S5**). Bi-weekly active (outbound phone contact) and passive surveillance for symptoms of suspected Covid-19 illness began on Day 8 and continues through the end of the study (**Table S6; Figure S2**). A new onset of suspected symptoms of Covid-19 triggered initial and follow-up surveillance visits to perform clinical assessments (vital signs, including pulse oximetry, and a lung examination) and for collection of nasal swabs (**Figure S3**). In addition, suspected Covid-19 symptoms were also queried, and nasal swabs collected, at all scheduled study visits. Nasal swab samples were tested for the presence of SARS-CoV-2 by NAAT using the BD MAX™ system (Becton Dickinson). The InFLUenza Patient-Reported Outcome (FLU-PRO^©^) questionnaire was utilized to comprehensively assess symptoms for the first 10 days of a suspected Covid-19 illness episode.

### Whole Genome Sequencing

We performed post-hoc whole virus genome sequencing of nasal samples of all primary efficacy endpoints in a blinded fashion. Details of whole genome sequencing methods and phylogenetic analysis are provided in the Supplementary Appendix (**Figure S1**).

### Statistical Analysis

The safety analysis population included all participants who received at least one injection of NVX-CoV2373 or placebo, with participants analyzed according to the treatment actually received. Safety analyses were presented as numbers and percentages of participants with solicited local and systemic adverse events analyzed through 7 days after each vaccination, and unsolicited adverse events through Day 35.

The per-protocol efficacy analysis population (PP-EFF) included baseline seronegative (by anti-spike IgG) participants who received both injections of NVX-CoV2373 or placebo as assigned, had no evidence of SARS-CoV-2 infection (by NAAT or anti-spike IgG) within 7 days after the second vaccination (ie, before Day 28), and had no major protocol deviations affecting the primary efficacy outcome.

Vaccine efficacy (%) was defined as (1 – RR) × 100, where RR = relative risk of Covid-19 illness between NVX-CoV2373 and placebo. The official, event-driven efficacy analysis targeted a minimum number of 23 endpoints to provide approximately 90% power to detect vaccine efficacy of 80% based on an incidence rate of symptomatic Covid-19 of 2% to 6% in the placebo group. This analysis was carried out at an overall one-sided type I error rate of 0.025 for the single primary efficacy endpoint. The RR and its confidence interval (CI) were estimated using Poisson regression with robust error variance. Hypothesis testing of the primary efficacy endpoint was carried out against the null hypotheses: H0: vaccine efficacy ≤ 0%. The success criterion required rejection of the null hypothesis to demonstrate a statistically significant vaccine efficacy.

## RESULTS

### Participants

A total of 6324 participants at 16 sites in South Africa were screened from August 17, 2020 through November 25, 2020. A total of 4387 participants received at least one injection of NVX-CoV2373 (n=2199) or placebo (n=2188), with 4332 participants receiving both injections (**Figure 1**).

**Figure 1.**
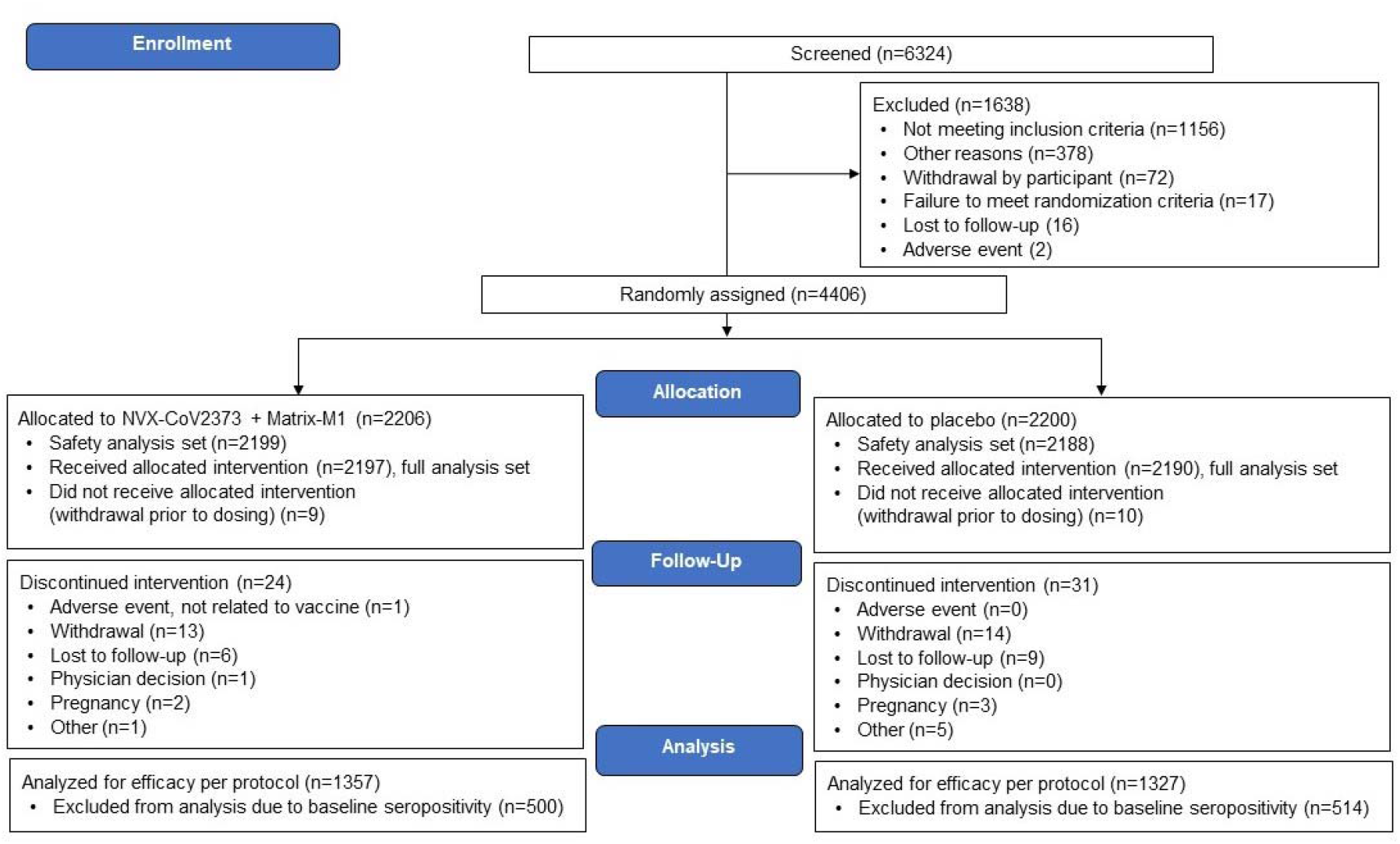
Disposition of Participants in the Trial. **Legend:** The full analysis set included all participants who were randomly assigned to treatment and received at least one dose, regardless of protocol violations or missing data, and are analyzed according to the trial vaccine group as randomized. This diagram represents the disposition of participants in the trial. The data cutoff date for the primary efficacy analysis was January 8, 2021, which represented a median follow-up of 66 and 45 days after first and second vaccination, respectively. The data cutoff date for the primary safety analysis was January 25, 2021, which included safety data through 35 days after first vaccination in all 968 Stage 1 participants (889 HIV-negative and 79 PLWH). The safety analysis population included all participants who received at least one dose of NVX-CoV2373 or placebo, with participants analyzed according to the treatment actually received. The per-protocol efficacy analysis population included baseline seronegative (by anti-spike IgG) participants who received both injection of NVX-CoV2373 or placebo as assigned, had no evidence of SARS-CoV-2 infection (by NAAT or anti-spike IgG) within 7 days after the second vaccination (ie, before Day 28), and had no major protocol deviations affecting the primary efficacy outcome. Abbreviations: HIV = human immunodeficiency virus; IgG = immunoglobulin G; NAAT = nucleic acid amplification test; PLWH = people living with HIV; SARS-CoV-2 = severe acute respiratory syndrome coronavirus 2.

Demographic and baseline characteristics were balanced (**Table 1**). The mean age of all participants was 32.0 years, and approximately 4% in each group were 65 to 84 years of age. Approximately 57% of the participants were male, and most were Black-African (95%). Twenty percent of participants were obese, 5.6% had hypertension, and 1.6% had type 2 diabetes. Approximately 30% of participants were seropositive at baseline by anti-S IgG antibodies (sensitivity 94.7% and specificity 96.4% at a predefined anti-S IgG threshold; see Supplementary Appendix).

**Table 1.** Demographics and Baseline Characteristics of the Participants Included in the Safety Analysis Set. **Table Legend** NVX-CoV2373 was 5 µg recombinant spike protein with 50 µg Matrix-M1. BMI was calculated as weight (kg) divided by squared height (m). SARS-CoV-2 serostatus was determined by IgG antibody to recombinant spike protein (anti-spike IgG). Baseline serostatus was defined by antibody level detected by anti-spike IgG ELISA using GMT at Day 0. Percentages were based on safety analysis set within each treatment and overall. Note: Values are represented as n (%), unless otherwise stated.

| Vaccine Group | NVX-CoV2373 | Placebo | Overall |
| --- | --- | --- | --- |
| N | 2199 | 2188 | 4387 |
| **Age (years)** |  |  |  |
| n | 2196 | 2186 | 4382 |
| Mean (SD) | 31.9 (12.91) | 32.2 (13.08) | 32.0 (13.00) |
| Median | 28.0 | 28.0 | 28.0 |
| **Age Group** |  |  |  |
| ≥18 to 64 years | 2104 (95.7) | 2094 (95.7) | 4198 (95.7) |
| ≥65 to 84 years | 92 (4.2) | 92 (4.2) | 184 (4.2) |
| **Sex** |  |  |  |
| Male | 1252 (56.9) | 1266 (57.9) | 2518 (57.4) |
| Female | 947 (43.1) | 922 (42.1) | 1869 (42.6) |
| **Race** |  |  |  |
| Black | 2098 (95.4) | 2082 (95.2) | 4180 (95.3) |
| White | 86 (3.9) | 66 (3.0) | 152 (3.5) |
| Other | 40 (1.8) | 49 (2.2) | 89 (2.0) |
| **Baseline BMI (kg/m<sup>2</sup>)** |  |  |  |
| n | 2195 | 2186 | 4381 |
| Mean (SD) | 25.06 (6.004) | 25.02 (5.930) | 25.04 (5.967) |
| ≥30 to 40 | 451 (20.5) | 440 (20.1) | 891 (20.3) |
| **Underlying Chronic Conditions** |  |  |  |
| Hypertension | 125 (5.7) | 119 (5.4) | 244 (5.6) |
| Type 2 diabetes | 31 (1.4) | 39 (1.8) | 70 (1.6) |
| **Day 0 SARS-CoV-2 NAAT Positive** | 63 (2.9) | 63 (2.9) | 126 (2.9) |
| **Day 0 SARS-CoV-2 Anti-Spike IgG Seropositive** | 651 (29.6) | 673 (30.8) | 1324 (30.2) |

Abbreviations: BMI = body mass index; ELISA = enzyme-linked immunosorbent assay; GMT = geometric mean titer; IgG = immunoglobulin G; NAAT = nucleic acid amplification test; SARS-CoV-2 = severe acute respiratory syndrome coronavirus 2; SD = standard deviation.

### Safety

Preliminary safety data were available on all Stage 1 participants, comprised of 968 participants, at the time of the primary efficacy analysis, with safety presented by baseline SARS-CoV-2 serostatus (665 seronegative and 303 seropositive) (Supplementary Appendix; **Table S8**). Briefly, solicited local and systemic adverse events were predominantly mild to moderate and transient, and more common in NVX-CoV2373 recipients. Injection site pain was the most frequently reported local solicited adverse event (37□39% and 15□16% in NVX-CoV2373 and placebo recipients, respectively, post-first dose) **(Table S9**); post-first and second dose rates were similar overall, with mean duration slightly higher after the second dose but generally less than 3 days. Severe local adverse events were infrequent but occurred more often in the seronegative NVX-CoV2373 group after the second dose (4%) versus placebo (1%). In NVX-CoV2373 recipients, the most common solicited systemic adverse events post-first and second dose were headache (20□25%), muscle pain (17□20%), and fatigue (12□16%). Post-first and second dose rates were similar overall, with mean duration slightly higher after the second dose ut generally less than 3 days. Severe systemic adverse events, albeit infrequent, increased in the seronegative NVX-CoV2373 group after the second dose (4%) versus placebo (2%), particularly fatigue and headache (**Tables S8 and S9**). Reactogenicity was generally similar in seronegative vs seropositive NVX-CoV2373 recipients.

Medically attended adverse events (**Table S10**) and serious adverse events (**Table S11**) were infrequent but occurred slightly more often in the NVX-CoV2373 group, with no apparent clustering of specific adverse events by treatment group, preferred term, or system organ class. To date, no serious adverse events have been assessed as related to trial vaccine by study investigators (**Table S8**). No prespecified vaccination pause rules were triggered.

### Efficacy

In the 2684 baseline seronegative participants (94% HIV-uninfected and 6% PLWH), evaluable for the primary efficacy analysis, 15 and 29 cases of symptomatic Covid-19 were observed after Day 28 among NVX-CoV2373 and placebo recipients, respectively, corresponding to vaccine efficacy of 49.4% (95% CI: 6.1 to 72.8), thereby meeting the primary efficacy endpoint success criterion (**Table 2**; **Figure 2A**). All of the per-protocol cases were mild to moderate Covid-19, except for one severe case in the placebo group.

**Table 2.** Vaccine Efficacy Against Symptomatic Covid-19 at Least 7 Days After the Second Dose (Day 28)

| Population/Baseline Anti-Spike IgG Serostatus | No. of Cases | NVX-CoV2373* |  | Placebo |  | VE (95% CI) |
| --- | --- | --- | --- | --- | --- | --- |
|  |  | n/N (%)† | (95% CI) | n/N (%)† | (95% CI) |  |
| All Participants |  |  |  |  |  |  |
| Baseline seronegative | 44 | 15/1357 (1.1) | (0.6, 1.8) | 29/1327 (2.2) | (1.5, 3.1) | 49.4%‡ (6.1, 72.8) |
| Baseline seropositive | 19 | 6/500 (1.2) | (0.4, 2.6) | 13/514 (2.5) | (1.4, 4.3) | 52.6% (-23.8, 81.8) |
| Regardless of baseline serostatus | 63 | 21/1857 (1.1) | (0.7, 1.7) | 42/1841 (2.3) | (1.6, 3.1) | 50.4% (16.6, 70.5) |
| Participants Without HIV |  |  |  |  |  |  |
| Baseline seronegative | 38 | 11/1281 (0.90) | (0.43, 1.5) | 27/1255 (2.2) | (1.4, 3.1) | 60.1% (19.9, 80.1) |
| Baseline seropositive | 19 | 6/467 (1.29) | (0.47, 2.8) | 13/484 (2.7) | (1.4, 4.5) | 52.2% (-24.8, 81.7) |
| Regardless of baseline serostatus | 57 | 17/1748 (0.97) | (0.57, 1.6) | 40/1739 (2.3) | (1.6, 3.1) | 57.7% (25.7, 75.9) |

Abbreviations: CI = confidence interval; Covid-19 = coronavirus 2 disease 2019; HIV = human immunodeficiency virus; N = number of participants; n = number of participants
with NAAT-confirmed Covid-19; NAAT = nucleic acid amplification test; PP-EFF = per-protocol efficacy; VE = vaccine efficacy.
\*Includes 50 µg Matrix-M1.
†Percentage of participants with Covid-19 calculated as $n/N \times 100$ .
‡Primary endpoint.
The 95% CI for PCR-confirmed Covid-19 was calculated using the exact Clopper-Pearson method. Participants were counted once if the participant reported one or more PCR-confirmed illness episodes. Log-linear model of NAAT-confirmed Covid-19 infection incidence rate using Poisson regression with treatment group as fixed effects and robust error variance.<sup>25</sup> $VE = 100 \times (1 - \text{Relative Risk})$ . Data shown are for the PP-EFF analysis set.

**Figure 2.**
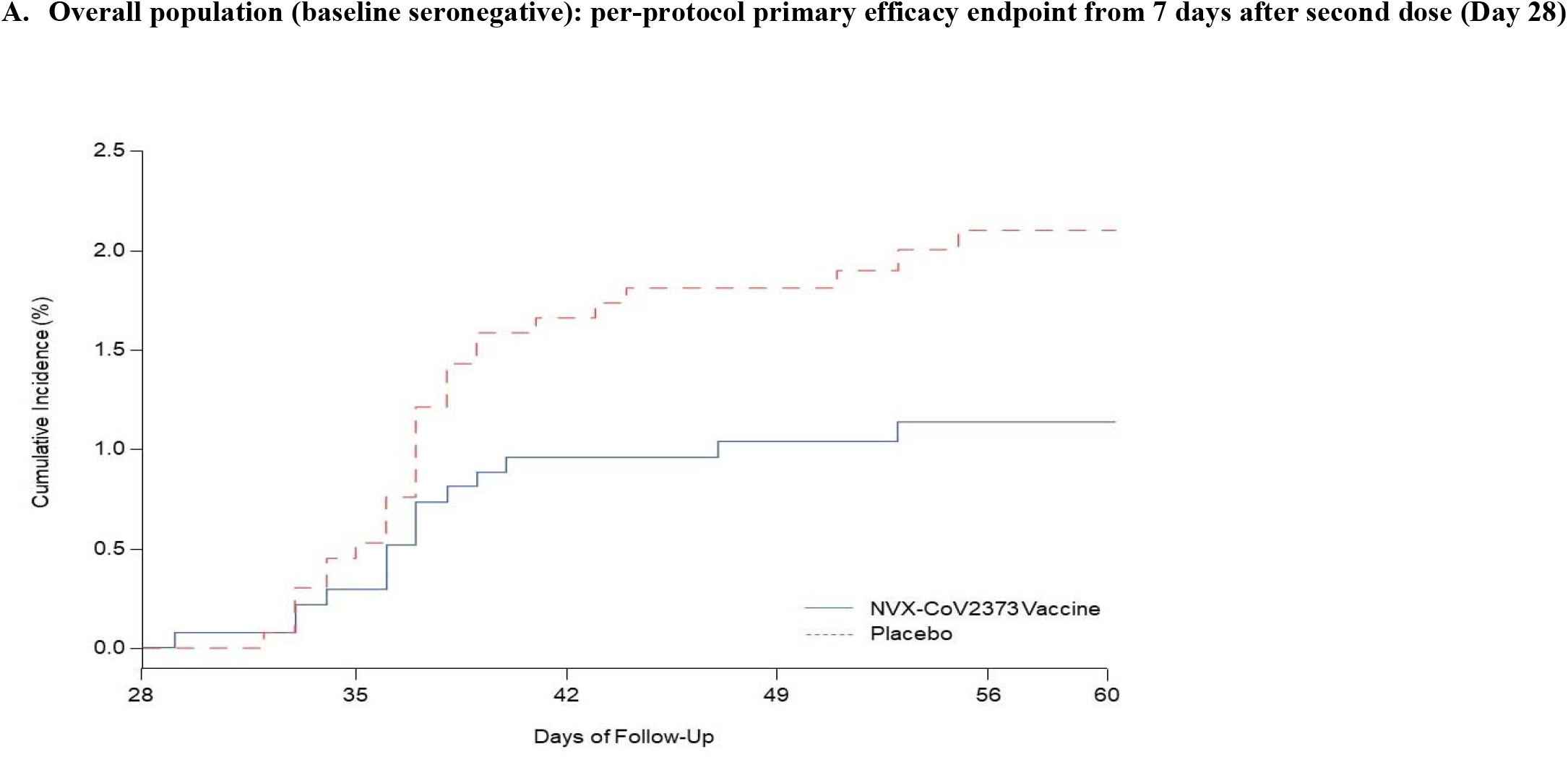
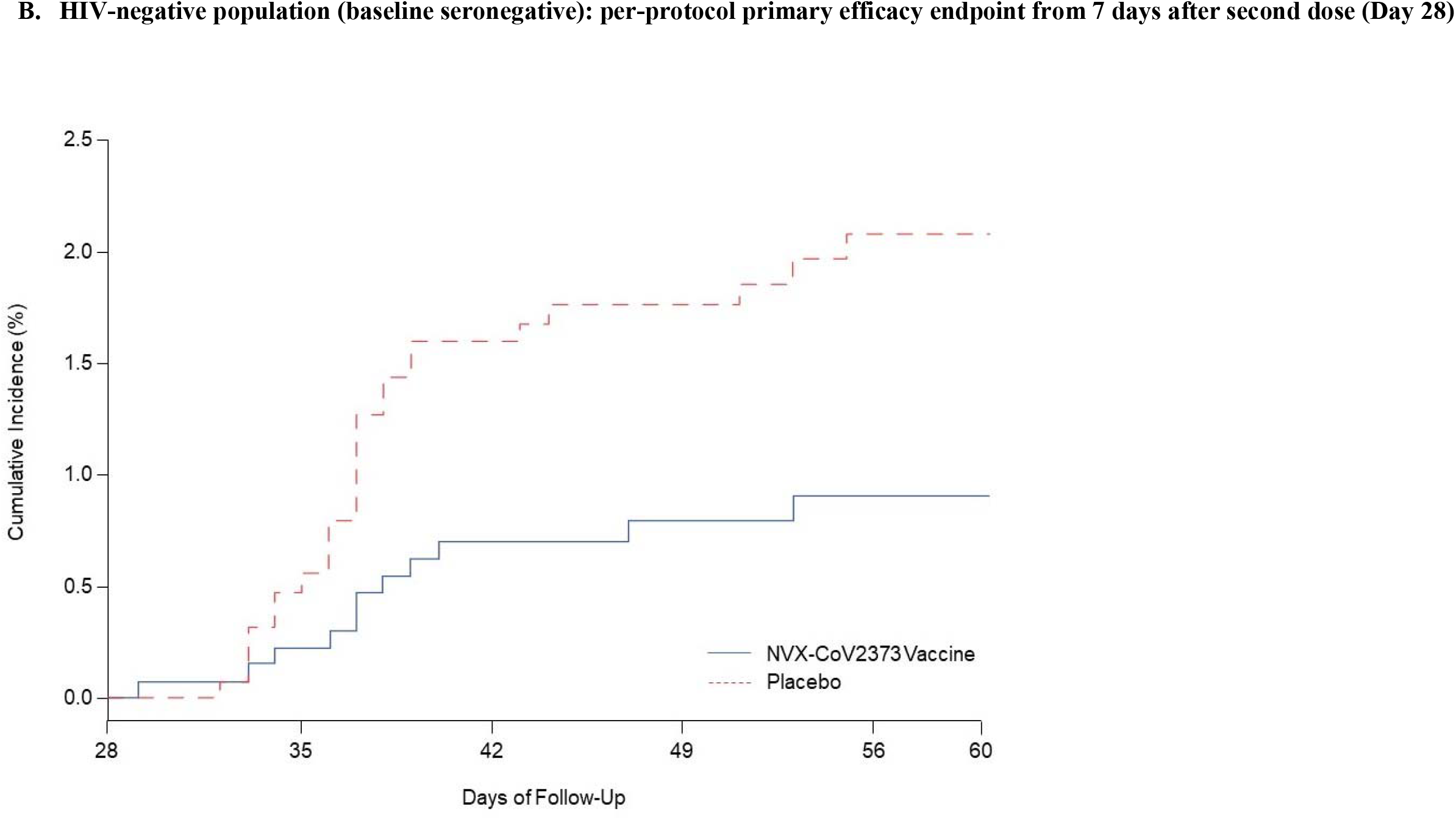
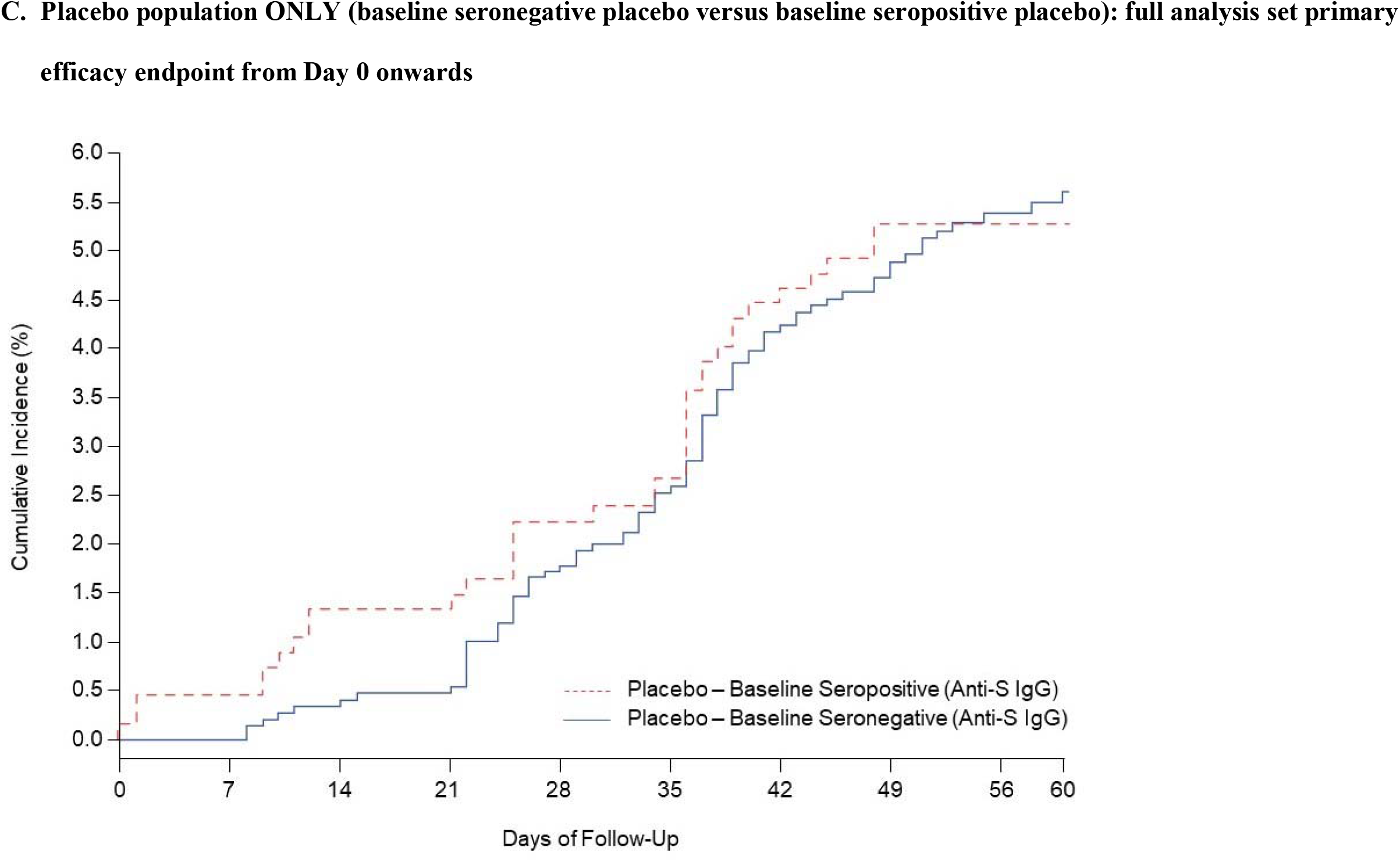
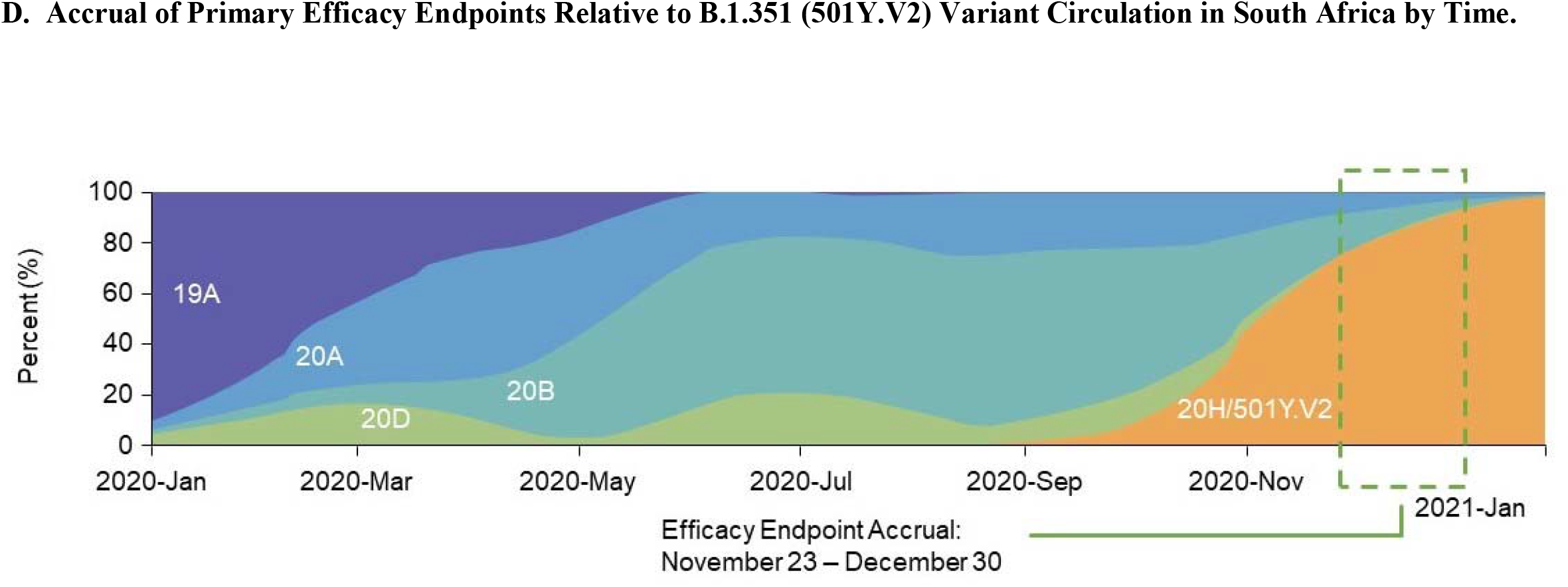
Kaplan-Meyer Plots of Efficacy of NVX-CoV2373 Against Symptomatic Covid-19, Risk of Symptomatic Covid-19 in Seropositive versus Seronegative Placebo Recipients, and Timing of Endpoint Accrual. **Legend:** The time period for surveillance of per-protocol symptomatic Covid-19 cases was from at least 7 days after the second dose (ie, Day 28) of NVX-CoV2373 or placebo through the first 2 months of the surveillance period (follow-up). Data shown are for the per-protocol efficacy population, unless otherwise indicated. A) Serologically naïve participants overall; B) Serologically naïve HIV-negative participants C) Placebo participants (serologically negative vs serologically positive) in the full analysis set (FAS) from Day 0 onwards. The FAS included all participants who were randomly assigned to treatment and receive at least 1 dose, regardless of protocol violations or missing data D) Per protocol efficacy endpoint accrual relative to distribution of variant as reported in Nextstrain.org. Source: Nextstrain.org. Freely available under the terms of the GNU Affero General Public License.

Among HIV-negative, baseline seronegative participants, 11 and 27 cases of symptomatic Covid-19 were observed among NVX-CoV2373 and placebo recipients, respectively, corresponding to vaccine efficacy of 60.1% (95% CI: 19.9 to 80.1) (**Table 2**; **Figure 2B**). Among adults without HIV, vaccine efficacy was unaffected by baseline SARS-CoV-2 serostatus.

Among baseline seronegative PLWH, there were four and two cases of symptomatic Covid-19 among NVX-CoV2373 and placebo recipients, respectively (N<109 in each group). No cases were observed in the baseline seropositive PLWH population (N<33 in each group). The 44 primary efficacy endpoint cases contributing to the analysis accrued between November 23 and December 30, 2020. Of these, 41 (93.2%) had whole genome sequence data available (samples from three cases in the placebo group could not be sequenced), and 38 (92.7%) of 41 were identified as the B.1.351 variant, thereby mirroring the national time trend in circulation of the variant during the same period (**Figure 2D; Figure S1**). In a post-hoc analysis, vaccine efficacy against the B.1.351 variant was 51.0% (95% CI: −0.6 to 76.2) in HIV-negative participants (11 cases in NVX-CoV2373 and 22 in placebo recipients) and 43.0% (95% CI: −9.8 to 70.4) in the combined HIV-negative and PLWH population (14 cases in NVX-CoV2373 and 24 in placebo recipients).

Notably, during the first 60 days of follow-up, the incidence of Covid-19 observed in baseline seronegative placebo participants (5.3% [95% CI: 4.3 to 6.6]; 33 mild and 47 moderate cases out of 1516 participants) was comparable to the incidence in baseline seropositive placebo participants (5.2% [95% CI: 3.6 to 7.2]; 14 mild and 21 moderate cases out of 674 participants), suggesting that prior infection, presumptively with Wuhan-like prototype SARS-CoV-2, did not reduce the risk of subsequent Covid-19 illness likely due to the B.1.351 variant (**Figure 2C**).

## DISCUSSION

We describe preliminary evidence of the efficacy of a two-dose regimen of NVX-CoV2373 nanoparticle vaccine in preventing symptomatic Covid-19 in the setting of predominant transmission of the B.1.351 variant in South Africa.^11,14^ The vaccine fulfilled the primary efficacy objective, demonstrating statistically significant vaccine efficacy of 49.4% in the combined HIV-negative and PLWH baseline seronegative study population. Among 94% of participants without HIV, vaccine efficacy was 60.1%. The study was not powered to detect efficacy in the small population of PLWH. Preliminary safety data continued to indicate an acceptable safety and reactogenicity profile.^10^

This is the first report to provide evidence, in the setting of a controlled vaccine trial, that prior infection with first-wave prototype-like, pre-B.1.351 viruses did not reduce the risk of Covid-19 due to re-infection with B.1.351 variants among placebo recipients. This finding has significant public health implications for pandemic modeling, control strategies, and vaccine development and deployment efforts. This observation is consistent with the lack of incremental benefit conferred by pre-existing immunity in vaccine recipients as evidenced by consistent levels of efficacy regardless of baseline serostatus. Although these findings require further confirmation, our observations suggest that vaccination with prototype-sequenced NVX-CoV2373 conferred cross-protection against an immunological escape variant, in stark contrast to naturally acquired immunity.

Intense transmission during the first wave in South Africa, high levels of resulting population immunity to prototype-like viruses (as observed in our study and corroborated in serosurveys^19^), and conditions sustaining a high force of infection in advance of the second wave, may have created a milieu favorable to the emergence of the B.1.351 variant. The B.1.351 variant is reported to have emerged in the Eastern Cape Province, South Africa, in October 2020, and rapidly spread to become the dominant circulating strain throughout the country during November and December 2020,^14^ coincident with the surge of second-wave transmission nationally. Our study, with sites dispersed across the country, accrued 44 cases of symptomatic Covid-19 contributing to the primary efficacy analysis between November 23 and December 30, 2020. Sequencing of nasal samples from primary efficacy endpoint cases confirmed a pattern consistent with national molecular epidemiology.

The B.1.351 variant is characterized by three deleterious mutations at key antigenic sites in the RBD, including N501Y, K417N, and E484K, with the latter two having particular functional impact.^12,14,16,17^ The N501Y mutation is known to increase binding affinity of the spike protein to the human angiotensin-converting enzyme 2 receptor,^20^ and has been reported to increase transmissibility of the B.1.17 variant circulating in the UK.^15^ The E484K mutation has been reported to abolish or substantially reduce neutralization by multiple potent monoclonal antibodies and polyclonal convalescent sera in both wild-type and pseudo-virus neutralization assays.^11,12,16,17,21^ Additionally, post-vaccination sera derived from volunteers receiving either of the mRNA vaccines showed 6.5-to 8.6-fold reductions in neutralizing capacity to the B.1.351 variant relative to prototype virus in pseudovirus neutralization^16^; however, the impact on clinical efficacy remains unassessed. Wild-type and pseudo-virus neutralization assays assessing the impact of the B.1.351 variant on the neutralizing capacity of NVX-CoV2373 vaccine-elicited antibodies are in progress. Nevertheless, this is the first study to provide definitive clinical evidence of cross-protection against antigenically drifted viruses. In the interim analysis of our UK phase 3 study, relatively high levels of efficacy were observed against both the matched prototype-like, pre-variant strains (vaccine efficacy 96%), and the B.1.1.7 variant (vaccine efficacy 86%).^22^ The high vaccine efficacy against the B.1.1.7 variant is consistent with the expected limited impact of the characteristic N501Y mutation (without a concomitant E484K mutation) on in vitro neutralization capacity of convalescent sera derived from prototype-like virus infections.^12,16^

Two other trials, partially or wholly conducted in South Africa and contemporaneous with circulation of the B.1.351 variant, have recently reported efficacy results. In the South African arm (N=6576) of a large multi-national phase 3 study evaluating the efficacy of a single dose of Ad26.COV2.S, preliminary vaccine efficacy against moderate to severe Covid-19 was reported to be 56%, with 95% of cases reportedly due to the B.1.351 variant; however, vaccine efficacy against all-severity Covid-19 specific to the B.1.351 variant has not yet been reported, precluding a direct comparison.^23^ In the second trial, ChAdOx1-nCoV19 was evaluated in a phase 2 trial (N=2026) in South Africa, in a study population resembling ours with predominantly mild to moderate Covid-19, and reported vaccine efficacy of 22% (95% CI: −50 to 60) overall, and 10% (95% CI: −77 to 55) against the B.1.351 variant, with the latter comprising 95% of cases.^24^ Our study was subject to certain limitations. The efficacy results are preliminary (median follow-up of 66 and 45 days following first and second doses, respectively), and are limited in scope to the primary endpoint and subgroups of the primary endpoint, as well as post-hoc analysis of B.1.351 variant sequencing data; therefore, caution is warranted in the interpretation of our results, particularly in the PLWH cohort, which represents a relatively small fraction of the study population. Importantly, at the time of analysis, the study had captured almost exclusively mild to moderate Covid-19 endpoints in a predominantly young, healthy population; consequently, we have not as yet been able to report on vaccine efficacy against severe Covid-19. Most large Covid-19 vaccine efficacy trials have observed increasing efficacy with longer follow-up periods than ours and have reported notably increased vaccine efficacy against severe vs mild to moderate disease.^4-7^ We may observe a similar pattern with additional follow-up and higher numbers of efficacy endpoints that are expected to accrue. Secondary efficacy analyses will shed light on efficacy against severe disease, and whether naturally acquired immunity to prototype-like virus modulates severity of infection due to variant viruses. In conclusion, we have demonstrated that a prototype-sequenced NVX-CoV2373 vaccine was efficacious and induced notable cross-protection in the setting of dominant circulation of B.1.351 variants that appear to have largely evaded natural immunity to prototype-like virus. Although cross-protection was clearly present at clinically meaningful levels, our observations support the rapid development of vaccine formulations containing the B.1.351 variant.

Supported by Novavax, Inc., The Bill and Melinda Gates Foundation, and the Coalition for Epidemic Preparedness Innovations.

Disclosure forms provided by the authors are available with the full text of this article at NEJM.org.

A data sharing statement provided by the authors is available with the full text of this article at NEJM.org.

We thank the participants who volunteered for this trial, and the members of the independent safety monitoring committee for their oversight and critical and timely review of the trial data. We also acknowledge the contribution of the 2019nCoV-501 Study Group (see the Supplementary Appendix); Mary Ward, Dina Fazio, Jennifer Lee, and Kathleen Gandarillas (Phase Five Communications) for editorial support funded by Novavax; and the PPD staff.

## Data Availability

These are interim data, and individual participants remain masked to individual vaccine assignment. Therefore, it would be inappropriate to share individual level results at this time.

https://clinicaltrials.gov/ct2/show/NCT04533399?term=NVX-CoV2373+Covid-19+Vaccine&draw=2&rank=2

http://www.novavax.com

