## Supplemental Text, Figures and Tables for "Preliminary Efficacy of the NVX-CoV2373 Covid-19 Vaccine Against the B.1.351 Variant"

**Supplementary Appendix**

This appendix has been provided by the authors to give readers additional information about their work.

Supplement to: Shinde V et al. Preliminary Efficacy of NVX-CoV2373 Covid-19 Vaccine Against the B.1.351 Variant. N Engl J Med. DOI:

Supplementary Appendix to Manuscript Entitled

**Preliminary Efficacy of NVX-CoV2373 Covid-19 Vaccine Against the B.1.351 Variant**

### 2019nCoV-501 Study Group

(PubMed listed, and ordered alphabetically by author name)

| **2019nCoV-501 Study Group Author** | **Affiliation** | **Location** |
| --- | --- | --- |
| Nasreen Abrahams | Wits Reproductive Health and HIV Institute, University of the Witwatersrand | Johannesburg, South Africa |
| Khatija Ahmed, MBChB | Setshaba Research Centre | Tshwane, South Africa |
| Gary Albert, MS | Novavax, Inc. | Gaithersburg, MD, USA |
| Moherndran Archary, PhD | Paediatric Infectious Diseases Unit, University of KwaZulu-Natal | Durban, South Africa |
| Vicky Baillie, PhD | South African Medical Research Council, Vaccines and Infectious Diseases Analytics Research Unit, Faculty of Health Sciences, University of the Witwatersrand | Johannesburg, South Africa |
| Gabriella Benadé, MBBS, MPH | Wits Reproductive Health and HIV Institute, University of the Witwatersrand | Johannesburg, South Africa |
| Chijioke Bennett, MD, MPH, MBA | Novavax, Inc. | Gaithersburg, MD, USA |
| Sutika Bhikha, MBBS | South African Medical Research Council, Vaccines and Infectious Diseases Analytics Research Unit, Faculty of Health Sciences, University of the Witwatersrand | Johannesburg, South Africa |
| As'ad Ebrahim Bhorat, MBBCh | Soweto Clinical Trials Centre | Johannesburg, South Africa |
| Qasim Bhorat, MBBCh, MSc | Soweto Clinical Trials Centre | Johannesburg, South Africa |
| Raziya Bobat | Respiratory and Critical Care Unit, Nelson R. Mandela School of Medicine, University of KwaZulu-Natal | Durban, South Africa |
| Michiel Andries Buys, MBChB | Limpopo Clinical Research Initiative | Rustenburg, North-West, South Africa |
| Nazira Carrim-Ganey, MBBCh | PEERMED Clinical Trial Centre | Kempton Park, South Africa |
| Sphiwe Cebekhulu | Paediatric Infectious Diseases Unit, University of KwaZulu-Natal | Durban, South Africa |
| Janice Chen | Novavax, Inc. | Gaithersburg, MD, USA |
| Iksung Cho, MS | Novavax, Inc. | Gaithersburg, MD, USA |
| Shane Cloney-Clark, BS | Novavax, Inc. | Gaithersburg, MD, USA |
| Tulio de Oliveira, PhD | Kwazulu-Natal Research Innovation and Sequencing Platform (KRISP), University of KwaZulu-Natal | Durban, South Africa |
| Chinar Desai | Novavax, Inc. | Gaithersburg, MD, USA |
| Alicia Desmond | Centre Aids Prevention Research South Africa (CAPRISA), University of KwaZulu-Natal | Durban, South Africa |
| Marthie de Villiers | Madibeng Centre for Research, Department of Family Medicine, School of Health, University of Pretoria | Pretoria, South Africa |
| Keertan Dheda, MBBCh | Centre for Lung Infection and Immunity, Division of Pulmonology, Department of Medicine and UCT Lung Institute, University of Cape Town | Cape Town, South Africa |
| Filip Dubovsky, MD, MPH | Novavax, Inc. | Gaithersburg, MD, USA |
| Amyneline Engelbrecht, MBChB | Madibeng Centre for Research, Department of Family Medicine, School of Health, University of Pretoria | Pretoria, South Africa |
| Aliasgar Esmail, MBChB | Centre for Lung Infection and Immunity, Division of Pulmonology, Department of Medicine and UCT Lung Institute, University of Cape Town | Cape Town, South Africa |
| Yakub Moosa Essack, MBBCH, MCFP | Soweto Clinical Trials Centre | Johannesburg, South Africa |
| Lee Fairlie, MBChB | Wits Reproductive Health and HIV Institute, University of the Witwatersrand | Johannesburg, South Africa |
| Emmanuel Faust, PhD | Novavax, Inc. | Gaithersburg, MD, USA |
| Jamie Fiske, MS | Novavax, Inc. | Gaithersburg, MD, USA |
| Leon Fouche, MBChB | Limpopo Clinical Research Initiative | Rustenburg, North-West, South Africa |
| Sharne Foulkes, MBChB | Joshua Research Centre | Bloemfontein, Free State, South Africa |
| Lou Fries, MD | Novavax, Inc. | Gaithersburg, MD, USA |
| Muhammed Ameen Fulat, MBChB | Limpopo Clinical Research Initiative | Rustenburg, North-West, South Africa |
| Zakir Gaffoor | South African TB Vaccine Initiative, University of Cape Town | Cape Town, South Africa |
| Nadia Gangat | Wits Reproductive Health and HIV Institute, University of the Witwatersrand | Johannesburg, South Africa |
| Hennie Geldenhuys, MBChB | South African TB Vaccine Initiative, University of Cape Town | Cape Town, South Africa |
| Greg M. Glenn, MD | Novavax, Inc. | Gaithersburg, MD, USA |
| Ameena Goga, MBChB | Health Systems Research Unit and HIV Prevention Research Unit, South African Medical Research Council | Cape Town, South Africa |
| Amina Goondiwala | Soweto Clinical Trials Centre | Johannesburg, South Africa |
| Vani Govender | Centre Aids Prevention Research South Africa (CAPRISA), University of KwaZulu-Natal | Durban, South Africa |
| Coert Grobbelaar, MBChB | Aurum Institute, University of Pretoria | Pretoria, South Africa |
| Tony Guiteau | Novavax, Inc. | Gaithersburg, MD, USA |
| Sherika Hanley, MBBS | Centre Aids Prevention Research South Africa (CAPRISA), University of KwaZulu-Natal | Durban, South Africa |
| Jose Heino-Kasbergen, MBChB | Madibeng Centre for Research, Department of Family Medicine, School of Health, University of Pretoria | Pretoria, South Africa |
| Zaheer Hoosain, MBChB | Joshua Research Centre | Bloemfontein, Free State, South Africa |
| Aylin Oommen Jose, MBChB | South African Medical Research Council, Vaccines and Infectious Diseases Analytics Research Unit, Faculty of Health Sciences, University of the Witwatersrand | Johannesburg, South Africa |
| Vandana Joshi | Novavax, Inc. | Gaithersburg, MD, USA |
| Dishiki Jenny Kalonji | South African TB Vaccine Initiative, University of Cape Town | Cape Town, South Africa |
| Akhona Kamalu | Respiratory and Critical Care Unit, Nelson R. Mandela School of Medicine, University of KwaZulu-Natal | Durban, South Africa |
| Faeeza Kana, MBBCH | Soweto Clinical Trials Centre | Johannesburg, South Africa |
| Margaret Kautz | Novavax, Inc. | Gaithersburg, MD, USA |
| Ntuthuko Khumalo | Centre Aids Prevention Research South Africa (CAPRISA), University of KwaZulu-Natal | Durban, South Africa |
| Girisha Kistnasami | South African TB Vaccine Initiative, University of Cape Town | Cape Town, South Africa |
| Anthonet Lombard Koen, MBChB | South African Medical Research Council, Vaccines and Infectious Diseases Analytics Research Unit, Faculty of Health Sciences, University of the Witwatersrand | Johannesburg, South Africa |
| Gertruida Kruger, MBChB | MERC Research | Middelburg, South Africa |
| Natasha Lalloo, MBChB | Respiratory and Critical Care Unit, Nelson R. Mandela School of Medicine, University of KwaZulu-Natal | Durban, South Africa |
| Umesh Lalloo, MBChB | Respiratory and Critical Care Unit, Nelson R. Mandela School of Medicine, University of KwaZulu-Natal | Durban, South Africa |
| Maggie Lewis, MS | Novavax, Inc. | Gaithersburg, MD, USA |
| Johan J. Lombaard, MBChB | Joshua Research Centre | Bloemfontein, Free State, South Africa |
| Cheryl Louw, MBChB | Madibeng Centre for Research, Department of Family Medicine, School of Health, University of Pretoria | Pretoria, South Africa |
| Angelique Luabeya, MBChB | South African TB Vaccine Initiative, University of Cape Town | Cape Town, South Africa |
| Shabir A. Madhi, MBBCh, PhD | South African Medical Research Council, Vaccines and Infectious Diseases Analytics Research Unit, Faculty of Health Sciences, University of the Witwatersrand | Johannesburg, South Africa |
| Edson Makambwa | Centre for Lung Infection and Immunity, Division of Pulmonology, Department of Medicine and UCT Lung Institute, University of Cape Town | Cape Town, South Africa |
| Mookho Malahleha | Setshaba Research Centre | Tshwane, South Africa |
| Mduduzi S. L. Masilela, MBChB | Setshaba Research Centre | Tshwane, South Africa |
| Lenah Masombuka | Madibeng Centre for Research, Department of Family Medicine, School of Health, University of Pretoria | Pretoria, South Africa |
| Michael J. Massare, PhD | Novavax, Inc. | Gaithersburg, MD, USA |
| Sajeeda Mawlana | Paediatric Infectious Diseases Unit, University of KwaZulu-Natal | Durban, South Africa |
| Linda Mbuthini | Centre for Lung Infection and Immunity, Division of Pulmonology, Department of Medicine and UCT Lung Institute, University of Cape Town | Cape Town, South Africa |
| Mbalenhle Mbuyisa | South African TB Vaccine Initiative, University of Cape Town | Cape Town, South Africa |
| Carey L. Medin, PhD | Novavax, Inc. | Gaithersburg, MD, USA |
| Simon Mendelsohn, MBChB | South African TB Vaccine Initiative, University of Cape Town | Cape Town, South Africa |
| Val Mikhailovski | Novavax, Inc. | Gaithersburg, MD, USA |
| Sibongile Mncube | Wits Reproductive Health and HIV Institute, University of the Witwatersrand | Johannesburg, South Africa |
| Rosie Mngqibisa, MBChB | Paediatric Infectious Diseases Unit, University of KwaZulu-Natal | Durban, South Africa |
| Mapule Moloi | Wits Reproductive Health and HIV Institute, University of the Witwatersrand | Johannesburg, South Africa |
| Dhayendre Moodley, PhD | Department of Obstetrics and Gynaecology, University of KwaZulu-Natal | Durban, South Africa |
| Nozibusiso Rejoice Mosia | Paediatric Infectious Diseases Unit, University of KwaZulu-Natal | Durban, South Africa |
| Lynelle Mottay | Centre for Lung Infection and Immunity, Division of Pulmonology, Department of Medicine and UCT Lung Institute, University of Cape Town | Cape Town, South Africa |
| Susan Neal, BS | Novavax, Inc. | Gaithersburg, MD, USA |
| Cathy Neate | Madibeng Centre for Research, Department of Family Medicine, School of Health, University of Pretoria | Pretoria, South Africa |
| Minenhle Ngcobo | Paediatric Infectious Diseases Unit, University of KwaZulu-Natal | Durban, South Africa |
| Thandeka Nkosi | Centre Aids Prevention Research South Africa (CAPRISA), University of KwaZulu-Natal | Durban, South Africa |
| Rashina Nundlal | Respiratory and Critical Care Unit, Nelson R. Mandela School of Medicine, University of KwaZulu-Natal | Durban, South Africa |
| Suzette Oelofse | Centre for Lung Infection and Immunity, Division of Pulmonology, Department of Medicine and UCT Lung Institute, University of Cape Town | Cape Town, South Africa |
| Faeezah Patel | Wits Reproductive Health and HIV Institute, University of the Witwatersrand | Johannesburg, South Africa |
| Nita Patel, MS | Novavax, Inc. | Gaithersburg, MD, USA |
| Rubeshan Perumal | Centre for Lung Infection and Immunity, Division of Pulmonology, Department of Medicine and UCT Lung Institute, University of Cape Town | Cape Town, South Africa |
| Sahir Yusuf Petkar, MBBCH | Soweto Clinical Trials Centre | Johannesburg, South Africa |
| Friedrich G. Petrick, MBChB | MERC Research | Middelburg, South Africa |
| Sundrapragasen Pillay, MBChB | Respiratory and Critical Care Unit, Nelson R. Mandela School of Medicine, University of KwaZulu-Natal | Durban, South Africa |
| Annah Pitsi, MBChB | Setshaba Research Centre | Tshwane, South Africa |
| Joyce S. Plested, PhD | Novavax, Inc. | Gaithersburg, MD, USA |
| Patricia Price-Abbott, RN | Novavax, Inc. | Gaithersburg, MD, USA |
| Patricia Reed | Novavax, Inc. | Gaithersburg, MD, USA |
| Andreana Robertson, MS | Novavax, Inc. | Gaithersburg, MD, USA |
| Justin Shenje, MBChB | South African TB Vaccine Initiative, University of Cape Town | Cape Town, South Africa |
| Vivek Shinde, MD, MPH | Novavax, Inc. | Gaithersburg, MD, USA |
| Nishanta Singh, MBChB | South African TB Vaccine Initiative, University of Cape Town | Cape Town, South Africa |
| Sheleika Singh | Respiratory and Critical Care Unit, Nelson R. Mandela School of Medicine, University of KwaZulu-Natal | Durban, South Africa |
| Gale Smith, PhD | Novavax, Inc. | Gaithersburg, MD, USA |
| Kathy Smith | Novavax, Inc. | Gaithersburg, MD, USA |
| Lourtacia Sokhela | Centre Aids Prevention Research South Africa (CAPRISA), University of KwaZulu-Natal | Durban, South Africa |
| Michele Tameris, MBChB | South African TB Vaccine Initiative, University of Cape Town | Cape Town, South Africa |
| Asha Thombrayil, MBChB | South African Medical Research Council, Vaccines and Infectious Diseases Analytics Research Unit, Faculty of Health Sciences, University of the Witwatersrand | Johannesburg, South Africa |
| Danellus van der Watt, MMed | Limpopo Clinical Research Initiative | Rustenburg, North-West, South Africa |
| Pieter-Louis Vollgraaff, MBChB | Limpopo Clinical Research Initiative | Rustenburg, North-West, South Africa |
| Nelisiwe Xaba | South African TB Vaccine Initiative, University of Cape Town | Cape Town, South Africa |
| Lu Yang | Novavax, Inc. | Gaithersburg, MD, USA |
| Jiu Zhao | Novavax, Inc. | Gaithersburg, MD, USA |
| Mingzhu Zhu, PhD | Novavax, Inc. | Gaithersburg, MD, USA |

### Supplemental Methods:

#### Eligibility Criteria

Participants at screening were either 1) Cohort 1: healthy, human immunodeficiency virus (HIV)-negative, male or female adults ≥18 to <85 years of age or 2) Cohort 2: medically stable, HIV-positive, male or female adults ≥18 to <65 years of age. Healthy status was determined by the investigator based on medical history, vital sign measurements, and physical examination at screening. Participants in Cohort 2 were medically stable at screening, as determined by the investigator (based on review of health status, vital signs, medical history, and targeted physical examination), were completely free of opportunistic infections in the 1 year prior to the first study vaccination, and vital signs were within normal ranges prior to the first vaccination. All participants gave informed consent prior to trial enrollment, and were able to attend trial visits, comply with trial requirements, and provide timely, reliable, and complete reports of adverse events.

Exclusionary criteria included any ongoing, symptomatic acute illness requiring medical or surgical care or chronic illness (excluding HIV in HIV-positive participants) or that required changes in medication in the past 2 months, indicating that chronic illness/disease was not stable (at the discretion of the investigator), acute respiratory and/or non-respiratory illness consistent with potential coronavirus disease 2019 (Covid-19) concurrent or within 14 days prior to first study vaccination (medical history and/or physical examination) or documented temperature of >38°C during this period, participation in research involving an investigational product (drug/biologic/device) within 45 days prior to first study vaccination, prior receipt of investigational or approved Covid-19 vaccine at any time, history of a diagnosis of suspected or confirmed Covid-19, receipt of an influenza vaccination within 14 days prior to first study vaccination, or any other vaccine within 4 weeks prior to first study vaccination, or planned vaccination within 5 weeks after first study vaccination. Participants were also excluded if they had any autoimmune or immunodeficiency disease/condition (iatrogenic or congenital), excluding HIV in HIV-positive participants, chronic administration (defined as more than 14 continuous days) of immunosuppressant, systemic glucocorticosteroids, if they received other immune‑modifying drugs within 90 days prior to first study vaccination (an immunosuppressant dose of glucocorticoid was defined as a systemic dose ≥10 mg of prednisone per day or equivalent; the use of topical, inhaled, and nasal glucocorticoids was permitted), or if they received immunoglobulin, blood-derived products, or other immunosuppressant drugs within 90 days prior to first study vaccination (excluding highly active anti‑retroviral therapy in HIV‑positive participants), or had any condition that in the opinion of the investigator would pose a health risk to the participant if enrolled or could interfere with evaluation of the vaccine or interpretation of trial results (including neurologic or psychiatric conditions deemed likely to impair the quality of safety reporting).

#### Randomization and Stratification

This study comprised two participant populations: 1) healthy HIV-negative adult male or non-pregnant female participants (Cohort 1) and 2) medically stable HIV-positive adult male or non-pregnant female participants (Cohort 2). Participants were randomly assigned in a blinded manner using the centralized Interactive Response Technology according to pre-generated randomization schedules. Participants were randomized in a 1:1 ratio via block randomization to receive study vaccine (severe acute respiratory syndrome spike protein nanoparticle vaccine with Matrix-M1™ adjuvant [NVX‑CoV2373] or placebo).

#### Blinding

This was an observer-blinded study. To maintain the blind, placebo vaccination via the intramuscular route was included, and unblinded site personnel managed vaccine logistics, preparation, and administration (if necessary) so as to maintain the blind from the remainder of the site personnel and participants. The unblinded site personnel were not involved in study‑related assessments or had participant contact for data collection following study vaccine administration. Unblinding at the participant level for Novavax, except for selected study personnel (eg, safety physician), occurred at the time of the formal analysis of the primary efficacy endpoint. All study participants, site staff, and Contract Research Organization (CRO) staff continued to remain blinded at the participant level until End-of-Study, except for pre-designated unblinded study personnel at study site and CRO.

A participant’s treatment assignment was not broken until the end of the study for the clinical site study team unless medical treatment of the participant depended on knowing the study treatment the participant received. If the blind was broken because of a medical emergency, the investigator unblinded an individual participant’s treatment allocation.

#### Vaccination Pause Rules

Vaccination pause rules based on reactogenicity, adverse events, and serious adverse events (SAEs) related to study participation were in place to monitor participant safety during the study and govern advancement from Stage 1 to Stage 2 vaccination based on assessment by the Safety Monitoring Committee (SMC).

Adverse events meeting any one of the following criteria resulted in a hold being placed on subsequent vaccinations pending further review by the SMC and the sponsor:

- The occurrence of two or more related SAEs (final assessment by the sponsor per United States Food and Drug Administration [FDA] Center for Biologics Evaluation and Research Guidance) in a given *Medical Dictionary of Regulatory Affairs* (MedDRA) system organ class within the first 7 days (Day 7) following Stage 1 vaccination in each cohort.
- Any toxicity grade 3 (severe) solicited (local or systemic) single adverse event term occurring in ≥10% of participants in the NVX-CoV2373 group within the first 7 days (Day 7) following Stage 1 vaccination in each cohort.
- Any grade 3 (severe) unsolicited single adverse event preferred term for which the investigator assessed as related to study vaccine that occurred in ≥5% of participants in the NVX-CoV2373 group, within the first 7 days (Day 7) following Stage 1 vaccination in each cohort.

#### Ethics, Oversight, Trial Registration, and Funding

The trial was performed in accordance with the International Council for Harmonisation Good Clinical Practice guidelines.

Prior to initiation of a study site, approval was obtained from an Human Research Ethics Committee (HREC)/Institutional Review Board (IRB) before participation of human participants in research studies, as required by regulatory authority regulations and the International Council for Harmonisation of Technical Requirements for Pharmaceuticals for Human Use E6(R2) guidelines. Before study onset, the protocol, informed consent, advertisements used for the recruitment of study participants, and any other written information regarding this study provided to the participant were approved by the HREC/IRB. Documentation of all HREC/IRB approvals and of the HREC/IRB compliance with the ICH E6(R2) guidelines was maintained by the study site and was available for review by the sponsor or its designee.

All HREC/IRB approvals were signed by the HREC/IRB chairman or designee and identified the HREC/IRB name and address, the clinical protocol by title or protocol number or both, and the date approval or a favorable opinion was granted.

The trial was designed by Novavax, Inc. (Gaithersburg, Maryland, USA), with funding support from the Bill and Melinda Gates Foundation, and investigational vaccine manufacturing support from the Coalition for Epidemic Preparedness Innovations. The trial protocol was approved by the South African Health Products Regulatory Authority (SAHPRA; Ref 20200420) and Institutional Ethics Review Boards, and registered on Clinicaltrials.gov (NCT045333990 and the Pan African Clinical Trials Registry (PACTR202009726132275).

Safety oversight, including for specific vaccination pause rules, was performed by an independent Safety Monitoring Committee.

#### Trial Investigational Products

##### NVX-CoV2373

NVX-CoV2373 is constructed from the full-length wild-type SARS‑CoV-2 spike glycoprotein (GP) based upon the GenBank gene sequence MN908947, nucleotides 21563 to 25384,^1^ and co‑formulated with the saponin-based Matrix-M1 adjuvant. The recombinant spike protein is a type 1 trimeric GP of 1273 amino acids that is produced as an inactive S0 precursor^2^ and is comprised of a full‑length spike protein with amino acid substitutions in the S1/S2 cleavage domain furin cleavage site to confer protease resistance and also two proline substitutions (K986P and V987P) introduced in the heptad repeat 1 domain to produce a stable prefusion conformation. The gene for the recombinant spike protein, codon optimized for expression, was cloned using a baculovirus expression vector in *Spodoptera frugiperda* (Sf9) insect cells.^3,4^ The recombinant spike protein isolated from the Sf9 cell culture was solubilized and purified by several steps of chromatography and ultrafiltration. The saponin-based Matrix-M1 adjuvant has been shown to enhance the immunogenicity of nanoparticle vaccines in non-clinical and clinical studies. In non-clinical studies, NVX-CoV2373-immunized cynomolgus and rhesus macaques were protected against both upper and lower respiratory tract infection and pulmonary disease following intranasal and intratracheal challenge with SARS‑CoV-2.^5^

The following supplies were used for vaccination in the study:

| Product | Supplied Formulation |
| --- | --- |
| **Cohort 1: HIV-Negative Participants** | |
| NVX-CoV2373 | Solution for preparation for injection, at a concentration of 5 µg antigen and 50 µg adjuvant |
| Placebo | Sodium chloride injection (BP, sterile), 0.9% |
| **Cohort 2: HIV-Positive Participants** | |
| NVX-CoV2373 | Solution for preparation for injection, at a concentration of 5 µg antigen and 50 µg adjuvant |
| Placebo | Sodium chloride injection (BP, sterile), 0.9% |

Abbreviations: BP = British Pharmacopoeia; NVX-CoV2373 = severe acute respiratory syndrome coronavirus 2 recombinant spike protein nanoparticle vaccine co-formulated with Matrix-M1 adjuvant.

##### Matrix-M1 Adjuvant

Matrix-M1 is a saponin-based adjuvant, which is co-administered with an antigen to induce an enhanced immune response.^6^ The mechanism of Matrix-M1 is not well defined, but it has been associated with a potent induction of leukocyte activation and migration into the draining lymph nodes. Even though there is no clear evidence of a specific pattern-recognition receptor being stimulated by Matrix-M1, it has been shown to induce a strong T-cell response in multiple virus vaccine models.^7^ Matrix‑M1, and closely related adjuvants, administered with diverse vaccine antigen targets, have been shown to be antigen dose-sparing, enhance antigen presentation/cross-presentation, expand the epitope repertoire, induce cross-neutralizing antibody responses, and boost induction of polyfunctional CD4+ and CD8+ T-cell responses.^7,8-11^

Matrix-M1 is manufactured by mixing defined, partially purified extracts of the bark of the *Quillaja saponaria* Molina tree, termed Fraction-A and Fraction-C, with cholesterol and phosphatidylcholine in the presence of a detergent.^12^

Safety data for the Matrix-M1 adjuvant from over 4200 participants in clinical studies with Ebola virus glycoprotein (EBOV GP), respiratory syncytial virus F, malaria, rabies, herpes simplex virus, and influenza vaccines with Matrix‑M1, have been described.^13^

#### Trial Objectives and Endpoints

The trial objectives and endpoints described below are a subset of those that pertain to the results described in this manuscript. For a description of the full set of the trial objectives and endpoints, please refer to the clinical study protocol provided with the manuscript at NEJM.org.

##### Trial Objectives

The primary objectives of this trial were 1) to evaluate the efficacy of NVX-CoV2373 with Matrix‑M1 adjuvant compared to placebo on the occurrence of symptomatic mild, moderate, or severe confirmed Covid-19 as demonstrated by qualitative PCR in serologically naïve (to SARS‑CoV-2) healthy HIV‑negative and medically stable HIV-positive adult participants and 2) to accumulate and describe the safety experience for NVX-CoV2373 with Matrix‑M1 adjuvant based on solicited short-term reactogenicity across a broad age spectrum (by toxicity grade) for vaccination through Day 35 in healthy HIV-negative and medically stable HIV-positive adult participants regardless of baseline serostatus and stratified by baseline serostatus.

Secondary objectives of this trial included 1) evaluation of the efficacy of NVX-CoV2373 with Matrix-M1 adjuvant compared to placebo on the occurrence of symptomatic moderate or severe confirmed Covid-19 as demonstrated by qualitative PCR in serologically naïve (to SARS‑CoV‑2) healthy HIV‑negative and medically stable HIV-positive adult participants; 2) assessment of overall safety through Day 35 for all unsolicited adverse events and all medically attended adverse events [MAAEs], immunologically mediated adverse events of special interest (AESIs) or SAEs in healthy adult participants regardless of baseline serostatus and stratified by baseline serostatus.

##### Trial Endpoints

The primary endpoints of the trial were 1) positive (+) polymerase chain reaction (PCR)‑confirmed SARS‑CoV‑2 illness with symptomatic mild, moderate, or severe Covid‑19 in serologically naïve (to SARS-CoV-2) healthy HIV-negative and medically stable HIV-positive adult participants, analyzed overall, with a lower bound confidence interval (CI) of >0, from 7 days after the second vaccine dose (ie, Day 28) until the endpoint‑driven efficacy analysis was triggered by the occurrence of a prespecified number of blinded endpoints across the two study vaccine arms and/or at prespecified time points; 2) numbers and percentages (with 95% CIs) of participants with solicited adverse events (local, systemic) for 7 days following each vaccination (Days 0 and 21) by severity score, duration, and peak intensity in healthy HIV‑negative and medically stable HIV-positive adult participants, analyzed overall and separately, regardless of baseline serostatus and stratified by baseline serostatus. In the case of no toxicity, a score of zero (0) was applied; 3) numbers and percentages (with 95% CI) of participants with unsolicited adverse events (eg, treatment‑emergent, serious, suspected unexpected serious, AESIs, MAAEs) through Day 35 by MedDRA classification, severity score, and relatedness in HIV-negative and medically stable HIV-positive adult participants, analyzed overall and separately, regardless of baseline serostatus and stratified by baseline serostatus.

#### Statistics

##### Analysis Populations, Definitions

The intent-to-treat (ITT) analysis set included all participants who were randomly assigned to treatment, regardless of protocol violations or missing data. The ITT analysis sets were used for participant disposition summaries and analyzed according to the treatment arm in which the participant was randomly assigned to treatment.

The full analysis set included all participants who were randomly assigned to treatment and received at least one dose study vaccine (NVX-CoV2373 with Matrix-M1 adjuvant or placebo), regardless of protocol violations or missing data. The full analysis set was used as a supportive analysis population for the immunogenicity and efficacy analyses and analyzed according to the study vaccine group as randomized.

The safety analysis set included all participants who received at least 1 dose of study vaccine (NVX-CoV2373 with Matrix-M1 adjuvant or placebo). Data for participants in the safety analysis set were analyzed according to the vaccine actually received.

The per‑protocol efficacy (PP-EFF) analysis set included baseline seronegative participants who received both doses of study vaccine (NVX-CoV2373 with Matrix-M1 adjuvant or placebo) and had no major protocol deviations affecting the primary efficacy outcome as assessed by the sponsor prior to unblinding. All analyses of the PP‑EFF population excluded any illness episodes with positive SARS-CoV-2 by any validated PCR and/or serum antibody (anti‑nucleocapsid [anti-N] or anti-S) occurring before 7 days after the second dose (ie, Day 28). A second per‑protocol efficacy (PP‑EFF-2) analysis set was defined in a similar fashion except without the exclusion of baseline seropositives to allow for analysis of efficacy in an unselected population.

The per-protocol immunogenicity (PP-IMM) analysis set included participants who received both doses of study vaccine (NVX-CoV2373 with Matrix-M1 adjuvant or placebo), had at least a baseline and one serum sample result available after vaccination, were negative for hepatitis B virus and hepatitis C virus at baseline, and had no major protocol violations that were considered clinically relevant to impact immunogenicity response at the corresponding study visit as assessed by the sponsor prior to unblinding. For each visit, the SARS-CoV-2 unexposed population also excluded any illness episodes with positive SARS-CoV-2 by any validated PCR and/or serum antibody (anti-N or anti-S) if available, prior to each visit. Prior exposed participants were determined using baseline SARS‑CoV‑2 immunity defined as positive SARS‑CoV-2 by qualitative PCR and/or serum antibody (anti-N or anti-S) if available at baseline or positive SARS‑CoV-2 by qualitative PCR through Day 21 or Day 35, according to the specified analysis. Analysis was performed to assess if immune responses differed between exposed and unexposed individuals (ie, whether prior exposure altered dosing regimen considerations in a pandemic response).

The review and determination for exclusion from the per-protocol populations was carried out in a blinded fashion by a study clinician prior to unblinding for each interim evaluation based on all available information from the locked database.

##### Sample Size Justification

This study was designed to enroll a minimum of approximately 3200 to a maximum of approximately 4404 participants divided among two cohorts as follows:

- At least 2960 to 4164 participants ≥18 to <85 years of age that included a minimum of approximately 1480 participants who received NVX-CoV2373 with Matrix-M1 adjuvant
- 240 participants ≥18 to <65 years of age in Cohort 2 (HIV-positive) that included a minimum of approximately 120 participants who received NVX-CoV2373 with Matrix‑M1 adjuvant

The target sample size of 1480 actively immunized participants in Cohort 1 (HIV‑negative) was sufficient to detect an adverse event rate of at least 1 in 625 participants (ie, background rates of 0.16%) with 90% probability.

The target sample size of 120 actively immunized participants in Cohort 2 (HIV-positive) was sufficient to detect an adverse event rate of at least 1 in 53 participants (ie, background rates of 1.9%) with 90% probability.

This study was designed as an event-driven statistical analysis. Power calculations were performed using the two-sided 95% CI to determine the target number of events needed for the desired power and the minimum vaccine efficacy (VE) needed to reject the null hypotheses (H_0_:VE ≤0%). The minimum target number of 23 primary endpoints in the overall population (HIV‑negative and HIV-positive) was determined to provide approximately 90% power for the primary endpoint based on the following conservative assumptions.

- A mild, moderate, or severe Covid-19 incident rate of 2% to 6% in the placebo group and a VE of 80%
- 90% evaluability rate for the PP-EFF population (assuming 10% unevaluable due to attrition and/or baseline-seropositive participants)

The upper limit of the target number of 50 primary endpoints was chosen so that the observed minimum VE of ~65% would result in rejecting the H_0_:VE ≤30%.

Based on the above assumptions, the minimum target numbers of endpoints needed are 23 to 50 mild, moderate, or severe Covid-19 endpoints. The power calculations for the primary endpoint are summarized in the Table below.

| **Number of Mild, Moderate, or Severe Covid-19 Endpoints** | **Approximate Minimum VE Need to Reject H_0_: VE ≤0% (ie, LBCI >0%)** | **Approximate Minimum VE Need to Reject H_0_: VE ≤30% (ie, LBCI >30%)** |
| --- | --- | --- |
| 20 | 67% | 82% |
| 23 | 65% | 79% |
| 25 | 61% | 75% |
| 30 | 57% | 75% |
| 35 | 54% | 70% |
| 40 | 52% | 67% |
| 45 | 50% | 68% |
| 50 | 44% | 65% |
| 55 | 43% | 63% |
| 60 | 41% | 60% |

Abbreviation: CI = confidence interval; Covid-19 = coronavirus disease 2019; LBCI = lower bound of the confidence interval; VE = vaccine efficacy.

##### Efficacy Analysis

The single primary efficacy endpoint, mild, moderate, or severe COVID-19, was analyzed on the PP-EFF population. Conclusions concerning declaration of attainment of the primary efficacy endpoint at the completion of the study was only based on the PP-EFF population. In addition, supportive analyses based on the full analysis set population, further excluding subjects with positive SARS-CoV-2 by qualitative PCR between Day 0 and Day 6 (inclusive), was also performed.

The VE is defined as VE (%) = (1 – RR) × 100, where RR = relative risk of incidence rates between the two study vaccine groups (SARS-CoV-2 rS with Matrix-M1 adjuvant or placebo). The official (ie, event-driven) analysis for the primary objective in the PP-EFF population was carried out at an overall one-sided type I error rate of 0.025 for the single primary endpoint.

The RR and its CI was estimated using Poisson regression with robust error variance.^14^ The generalized linear model with unstructured correlation matrix (robust error variances) was used. The explanatory variables in the model included the study vaccine group. The dependent variable was the incidence rate of the endpoint of interest. The robust error variances was estimated using repeated statement and the subject identifier. Poisson distribution was used with a link function logarithm.

Hypothesis testing of the primary efficacy endpoint was carried out sequentially against the two prespecified null hypotheses: H_0_:VE ≤0% first and then H_0_:VE ≤30%. Rejection of the null hypothesis, H_0_:VE ≤0% demonstrates a statistically significant vaccine effect for the primary endpoint. If the primary endpoint meets the success criterion of the lower bound >0%, additional hierarchical testing of the primary endpoint against the more stringent success criterion of the lower bound >30% was to be carried out.

#### Immunologic Assay Method Details

SARS-CoV-2 Spike Protein Serum IgG ELISA (performed at Novavax Clinical Immune Laboratory (Gaithersburg, Maryland, USA). Recombinant SARS-CoV-2 (rSARS-CoV-2) S protein was immobilized onto the surface of the 96-well microtiter plate wells (100 µL per well) by direct adsorption for 15 to 72 hours at 2°C to 8°C at a concentration of 1 µg/mL in PBS as per P_SOP_02483 (validated method). Plates were washed four times with 300 µL/well PBST, blocked with 300 µL blocking buffer for 1 to 1.5 hours at 24°C ± 2°C. Diluted reference standard (2-fold dilution series of 12 dilutions starting 1:1000) and human serum samples (3-fold dilution series of 12 dilutions) in assay buffer (1% milk in PBS) starting at 1:100 dilution are then added in duplicate (100 µL per well) to the rSARS-CoV-2 S protein-coated wells and any specific antibodies are allowed to complex with the coated antigen for 2 hours ± 10 minutes at 24°C ± 2°C. Plates are washed six times with 300 µL/well PBST. Antibodies bound to the rSARS-CoV-2 S protein are then detected using a horseradish peroxidase (HRP) conjugate goat anti-human IgG antibody diluted 1: 2000 (Southern Biotech cat no. 2040-05) incubated for 1 hour ± 10 minutes at 24°C ± 2°C, washed three times with 300 µL/well PBST, and a colorimetric signal generated by addition of 100 µL per well 3, 3′,5,5′-tetramethylbenzidine (TMB) chromogenic substrate for 10 minutes ± 2 minutes at 24°C ± 2°C. After incubation was complete, the TMB reaction was stopped with 100 µL per well of TMB Stop solution. The absorbance was measured at 450 nm using a Molecular Device 96-well plate reader. When binding reagents (coated antigen and secondary antibody) are in excess, the optical density (OD) of the chromogenic substrate at endpoint is proportional to the quantity of anti-rSARS-CoV-2 S IgG present in the serum sample. The total anti-rSARS-CoV-2 S protein IgG antibody level in a serum sample was quantitated in ELISA unit, EU/mL, by comparison to a reference standard curve. The results were analyzed in singleton by SoftMax Pro software using 4-PL curve fit. Assay included control plates comprising of positive controls and negative control. The assay was qualified and validated prior to clinical trial testing, with the validation protocol addressing repeatability and intermediate precision, linearity of response, limits of detection and quantitation, specificity, and robustness.

#### Immunologic Assay Threshold for Prior SARS-CoV-2 Infection

An ad hoc analysis of existing anti-SARS-CoV-2 spike protein serum IgG antibody concentration data, developed by the Novavax Clinical Immunology laboratory per P_SOP_02428 (qualified method) and P_SOP_02483 (validated method) with rSARS-CoV-2 S protein as the coating antigen, was performed to establish a diagnostic threshold for the presence of prior SARS-CoV-2 infections because we needed a cutoff with high baseline seropositivity in South Africa samples.

This analysis included 301 known SARS-CoV-2 convalescent sera samples kindly provided by Baylor College of Medicine Molecular Virology Laboratory (Dr. P.A. Piedra) and commercial vendors as known positive samples. Also evaluated as unexposed/negative samples were 553 pre-COVID-19 serum samples; these were randomly selected baseline pre‑vaccination samples from clinical studies conducted between December 2010 and May 2018.

Considering a matrix of test results as follows:

| **Parameter** | | **Test Result** | |
| --- | --- | --- | --- |
|  |  | **Positive** | **Negative** |
| Known Past Disease | Positive | a | c |
|  | Negative | b | d |

a = number of test positive by the assay in question among the true positive samples (convalescent samples).

b = number of test positive sample by the assay in question among the true negative samples (Day 0 samples from the study).

c = number of test negative by the assay in question among the true positive samples (convalescent samples).

d = number of test negative sample by the assay in question among the true negative samples (Day 0 samples from the study).

Sensitivity is defined as the “true positive rate,” equivalent to a/a+c. Specificity is defined the “true negative rate,” equivalent to d/b+d. Positive predictive value (PPV) is defined as the proportion of people with a positive test result who actually had the infection, (a/a+b); negative predictive value (NPV) is defined as the proportion of those with a negative result who did not have the infection, (d/c+d).

The primary statistical measure used to determine the diagnostic threshold for the prior SARS‑CoV-2 infection was Youden’s J statistic calculated as Sensitivity + Specificity - 1 for each threshold level examined. For the current application, it was deemed desirable to identify for exclusion as many previously infected subjects as possible (sensitivity) to optimize the assessment of vaccine efficacy, while ensuring that the great majority of those excluded did indeed have prior infection (specificity) to avoid unnecessary loss of sample size. Youden’s J includes consideration of both sensitivity and specificity. Potential diagnostic threshold levels evaluated ranged from 200 (LLOQ) to 1500 EU/mL in increments of 100, with examinations of 50 EU/mL intervals near the peak. To further understand the performance characteristics of the proposed diagnostics threshold, PPVs and NPVs were generated using a resampling with replacement approach with 1000 simulation runs each for a sample size of 100. The unknown population prior COVID-19 infection rates evaluated were 5%, 10%, 20%, 30%, 40%, 50%, and 60%.

The following table summarizes the sensitivity and specificity for the several candidate threshold levels in ELISA unit (EU/mL). The threshold of 500 EU/mL exhibits the best overall performance characteristics with the highest observed Youden’s J statistic. It is the only threshold that simultaneously provides ~95% or higher sensitivity and specificity.

| **Threshold (≥)**  **EU/mL** | **Sensitivity (N=301)** | | **Specificity (N=553)** | | **Youden's J Statistics** |
| --- | --- | --- | --- | --- | --- |
|  | **Estimate** | **95% CI** | **Estimate** | **95% CI** |  |
| 200 (LLOQ) | 96.3% | (93.6, 98.2) | 84.3% | (81.0%, 87.2%) | 0.8061 |
| 300 | 95.3% | (92.3, 97.4) | 89.7% | (86.9%, 92.1%) | 0.8504 |
| 400 | 94.7% | (91.5, 96.9) | 93.7% | (91.3%, 95.6%) | 0.8836 |
| 450 | 94.7% | (91.5, 96.9) | 95.3% | (93.2%, 96.9%) | 0.8998 |
| **500** | **94.7%** | **(91.5, 96.9)** | **96.4%** | **(94.5%, 97.8%)** | **0.9107** |
| 550 | 94.0% | (90.7, 96.4) | 96.4% | (94.5%, 97.8%) | 0.9040 |
| 600 | 93.7% | (90.3, 96.2) | 96.6% | (94.7%, 97.9%) | 0.9025 |
| 700 | 93.0% | (89.5, 95.6) | 97.1% | (95.3%, 98.3%) | 0.9013 |
| 800 | 92.0% | (88.4, 94.8) | 97.1% | (95.3%, 98.3%) | 0.8913 |
| 900 | 91.0% | (87.2, 94.0) | 97.5% | (95.8%, 98.6%) | 0.8850 |
| 1000 | 90.4% | (86.5, 93.5) | 97.6% | (96.0%, 98.7%) | 0.8801 |
| 1100 | 90.0% | (86.1, 93.2) | 97.8% | (96.2%, 98.9%) | 0.8786 |
| 1200 | 89.4% | (85.3, 92.6) | 98.4% | (96.9%, 99.3%) | 0.8774 |
| 1300 | 88.4% | (84.2, 91.8) | 98.6% | (97.2%, 99.4%) | 0.8693 |
| 1400 | 87.7% | (83.5, 91.2) | 98.7% | (97.4%, 99.5%) | 0.8644 |
| 1500 | 87.0% | (82.7, 90.6) | 98.7% | (97.4%, 99.5%) | 0.8578 |

Abbreviations: CI = confidence interval; LLOQ = lower limit of quantification.

The following table summarizes the PPVs and NPVs for the threshold of 500. The threshold provided very reliable negative predictive value, ≥91.8%, across all background rates of prior SARS-CoV-2 infection rates up to 60% evaluated for this report, and >97% when background rates are ≤30%. Thus, less than 3% of subjects deemed negative based on this threshold in fact have had prior infection so long as the background population rate is ≤30%. Given that screening active infection rate by NAAT alone in various South African sites was between 20% and 30% during enrollment, the PPV should be not less 88-92%; implying that no more than 1 out of 7 to 11 subjects excluded from the per-protocol analysis set may not have had prior SARS-CoV-2 infection. This relatively low proportion is a desired characteristic of the intended use in a COVID-19 vaccine clinical trial setting where the goal is to minimize the exclusion of subjects who have not experienced prior SARS-CoV-2 infection from the primary analysis population (ie, per‑protocol analysis set).

| **Population Prior COVID-19 Infection Rate** | **Diagnostic Threshold (EU/mL)** | **Positive Predictive Value (PPV)** | **Negative Predictive Value (NPV)** |
| --- | --- | --- | --- |
| 5% | 500 | 62.8% | 99.7% |
| 10% | 500 | 77.2% | 99.3% |
| 20% | 500 | 88.1% | 98.5% |
| 30% | 500 | 92.6% | 97.4% |
| 40% | 500 | 95.1% | 96.1% |
| 50% | 500 | 96.7% | 94.4% |
| 60% | 500 | 97.8% | 91.8% |

Abbreviations: Covid-19 = coronavirus disease 2019.

#### Whole Genome Sequencing

Whole genome sequencing of SARS-CoV-2 positive samples were sequenced either at VIDA or the KwaZulu-Natal Research Innovation and Sequencing Platform (KRISP) using Superscript IV with random hexamers (Life Technologies, Carlsbad, California, USA) to generate cDNA. The Swift Biosciences’ Normalase Amplicon and SARS-CoV-2 amplicon panel^15^ or the ARCTIC V3 protocol^16-18^ and Illumina® Nextera Flex DNA Library Prep Kits were used to amplify, and index paired end libraries of genomic DNA. The resulting libraries were sequenced using either a 300 cycle v2 iSeq 100 Reagent Kit on an illumina iSeq 100 instrument or a 500 cycle v2 MiSeq Reagent Kit on an Illumina MiSeq instrument (Illumina, San Diego, California, USA).

#### Genome Assembly and Phylogenetic Analysis

Genome Detective 1.126 and the Coronavirus Typing Tool (https://www.genomedetective.com) were used to generate paired-end fastq reads.^19^ The bcftools 1.7-2 mpileup method was then used to filter out the low-quality mutations, and all the sequences were deposited in GISAID (https://www.gisaid.org/). A custom pipeline based on a local version of NextStrain was used for the phylogenetic analysis using the global reference dataset (N=2592). Genomes with low sequence genome coverage (<90%) were filtered out of the pipeline, which contains several python scripts that manage the analysis workflow. It performs alignment of genotypes in MAFFT, phylogenetic tree inference in IQ‑Tree20, tree dating and ancestral state construction and annotation (https://github.com/nextstrain/ncov). The new variant identified in South Africa is assigned the name 501Y.V2 and the corresponding PANGO lineage classification is B.1.351 (lineages version 2021-01-06).^20^

##### Figure S1. Phylogenetic Analysis of Whole Genome Sequencing of Viruses Isolated in Primary Endpoint Covid-19 Cases in South Africa

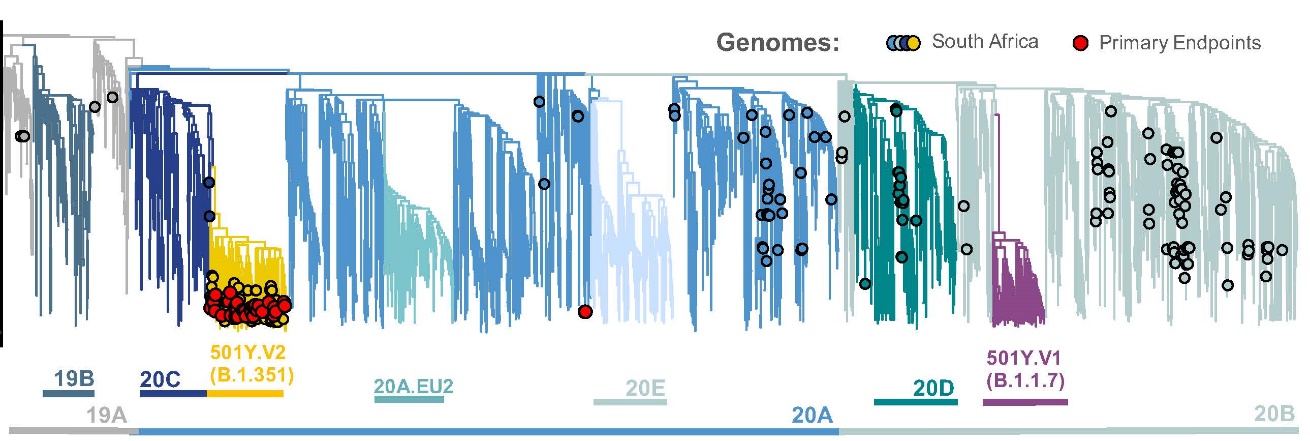

#### Adverse Events of Special Interest and Standard Toxicity Criteria

| Table S1. Potential Immune-Mediated Medical Conditions. | |
| --- | --- |
| **Categories** | **Diagnoses (as MedDRA* Preferred Terms)** |
| Neuroinflammatory Disorders: | Acute disseminated encephalomyelitis (including site specific variants: eg, non-infectious encephalitis, encephalomyelitis, myelitis, myeloradiculomyelitis), cranial nerve disorders including paralyses/paresis (eg, Bell’s palsy), generalized convulsion, Guillain-Barre syndrome (including Miller Fisher syndrome and other variants), immune‑mediated peripheral neuropathies and plexopathies (including chronic inflammatory demyelinating polyneuropathy, multifocal motor neuropathy and polyneuropathies associated with monoclonal gammopathy), myasthenia gravis, multiple sclerosis, narcolepsy, optic neuritis, transverse myelitis, uveitis |
| Musculoskeletal and Connective Tissue Disorders: | Antisynthetase syndrome, dermatomyositis, juvenile chronic arthritis (including Still’s disease), mixed connective tissue disorder, polymyalgia rheumatic, polymyositis, psoriatic arthropathy, relapsing polychondritis, rheumatoid arthritis, scleroderma (including diffuse systemic form and CREST syndrome), spondyloarthritis (including ankylosing spondylitis, reactive arthritis [Reiter's Syndrome] and undifferentiated spondyloarthritis), systemic lupus erythematosus, systemic sclerosis, Sjogren’s syndrome |
| Vasculitides: | Large vessels vasculitis (including giant cell arteritis such as Takayasu's arteritis and temporal arteritis), medium sized and/or small vessels vasculitis (including polyarteritis nodosa, Kawasaki's disease, microscopic polyangiitis, Wegener's granulomatosis, Churg–Strauss syndrome [allergic granulomatous angiitis], Buerger’s disease [thromboangiitis obliterans], necrotizing vasculitis and anti-neutrophil cytoplasmic antibody [ANCA] positive vasculitis [type unspecified], Henoch-Schonlein purpura, Behcet's syndrome, leukocytoclastic vasculitis) |
| Gastrointestinal Disorders: | Crohn’s disease, celiac disease, ulcerative colitis, ulcerative proctitis |
| Hepatic Disorders: | Autoimmune hepatitis, autoimmune cholangitis, primary sclerosing cholangitis, primary biliary cirrhosis |
| Renal Disorders: | Autoimmune glomerulonephritis (including IgA nephropathy, glomerulonephritis rapidly progressive, membranous glomerulonephritis, membranoproliferative glomerulonephritis, and mesangioproliferative glomerulonephritis |
| Cardiac Disorders: | Autoimmune myocarditis/cardiomyopathy |
| Skin Disorders: | Alopecia areata, psoriasis, vitiligo, Raynaud’s phenomenon, erythema nodosum, autoimmune bullous skin diseases (including pemphigus, pemphigoid and dermatitis herpetiformis), cutaneous lupus erythematosus, morphea, lichen planus, Stevens-Johnson syndrome, Sweet’s syndrome |
| Hematologic Disorders: | Autoimmune hemolytic anemia, autoimmune thrombocytopenia, antiphospholipid syndrome, thrombocytopenia |
| Metabolic Disorders: | Autoimmune thyroiditis, Grave’s or Basedow’s disease, Hashimoto thyroiditis,^†^ diabetes mellitus type 1, Addison’s disease |
| Other Disorders: | Goodpasture syndrome, idiopathic pulmonary fibrosis, pernicious anemia, sarcoidosis |
| ^*^ MedDRA denotes *Medical Dictionary for Regulatory Activities*.  † For Hashimoto thyroiditis: new onset only. | |

| Table S2. Adverse Events of Special Interest Relevant to COVID-19.^*^ | |
| --- | --- |
| **Body System** | **Diagnoses** |
| Immunologic | Enhanced disease following immunization, cytokine release syndrome related to COVID-19 disease,^†^ multisystem inflammatory syndrome in children (MIS-C) |
| Respiratory | Acute respiratory distress syndrome (ARDS) |
| Cardiac | Acute cardiac injury including:   - Microangiopathy - Heart failure and cardiogenic shock - Stress cardiomyopathy - Coronary artery disease - Arrhythmia - Myocarditis, pericarditis |
| Hematologic | Coagulation disorder   - Deep vein thrombosis - Pulmonary embolus - Cerebrovascular stroke - Limb ischemia - Hemorrhagic disease - Thrombotic complications |
| Renal | Acute kidney injury |
| Gastrointestinal | Liver injury |
| Neurologic | Guillain Barré syndrome, anosmia, ageusia, meningoencephalitis |
| Dermatologic | Chilblain-like lesions, single organ cutaneous vasculitis, erythema multiforme |
| ^*^ Covid-19 manifestations associated with more severe presentation and decompensation with consideration of enhanced disease potential. The current listing is based on Safety Platform for Emergency Vaccines (SPEAC) D2.3 Priority List of Adverse Events of Special Interest: COVID-19.^21^  † Cytokine release syndrome related to COVID-19 disease is a disorder characterized by nausea, headache, tachycardia, hypotension, rash, and/or shortness of breath.^22^  NOTE: AESI relevant to Covid-19 were reported only in association with a confirmed PCR-positive Covid-19 illness. | |

| Table S3. FDA Toxicity Grading Scales for Solicited Local and Systemic Adverse Events. | | | | |
| --- | --- | --- | --- | --- |
| Local Reaction to Injectable Product | Mild (Grade 1) | Moderate (Grade 2) | Severe (Grade 3) | Potentially Life Threatening (Grade 4) |
| Pain | Does not interfere with activity | Repeated use of nonnarcotic pain reliever >24 hours or interferes with activity | Any use of narcotic pain reliever or prevents daily activity | ER visit or hospitalization |
| Tenderness | Mild discomfort to touch | Discomfort with movement | Significant discomfort at rest | ER visit or hospitalization |
| Erythema/redness^*^ | 2.5 − 5 cm | 5.1 − 10 cm | >10 cm | Necrosis or exfoliative dermatitis |
| Induration/swelling^†^ | 2.5 − 5 cm and does not interfere with activity | 5.1 − 10 cm or interferes with activity | >10 cm or prevents daily activity | Necrosis |
| **Systemic (General)** | **Mild (Grade 1)** | **Moderate (Grade 2)** | **Severe (Grade 3)** | **Potentially Life Threatening (Grade 4)** |
| Fever^‡^ (°C)  (°F) | 38.0 − 38.4 100.4 − 101.1 | 38.5 − 38.9 101.2 − 102.0 | 39.0 − 40 102.1 − 104 | >40 >104 |
| Nausea/vomiting | No interference with activity or 1 − 2 episodes/24 hours | Some interference with activity or >2 episodes/24 hours | Prevents daily activity, or requires outpatient IV hydration | ER visit or hospitalization for hypotensive shock |
| Headache | No interference with activity | Repeated use of nonnarcotic pain reliever >24 hours or some interference with activity | Significant; any use of narcotic pain reliever or prevents daily activity | ER visit or hospitalization |
| Fatigue/Malaise^§^ | No interference with activity | Some interference with activity | Significant; prevents daily activity | ER visit or hospitalization |
| Myalgia | No interference with activity | Some interference with activity | Significant; prevents daily activity | ER visit or hospitalization |
| Arthralgia | No interference with activity | Some interference with activity | Significant; prevents daily activity | ER visit or hospitalization |
| ^*^ In addition to grading the measured local reaction at the greatest single diameter, the measurement should be recorded as a continuous variable.  ^†^ Induration/swelling should be evaluated and graded using the functional scale as well as the actual measurement.  ^‡^ Oral temperature; no recent hot or cold beverages.  ^§^ This event was captured separately as fatigue and malaise in the diary.  Source: DHHS 2007.^23^ | | | | |

| Table S4. FDA Toxicity Grading Scale for Vital Sign Abnormalities. | | | | |
| --- | --- | --- | --- | --- |
| Vital Sign Abnormalities | Mild (Grade 1) | Moderate (Grade 2) | Severe (Grade 3) | Potentially Life Threatening (Grade 4) |
| Tachycardia (bpm) | 101 – 115 | 116 – 130 | >130 | ER visit or hospitalization for arrhythmia |
| Bradycardia (bpm)* | 50 – 54 | 45 – 49 | <45 | ER visit or hospitalization for arrhythmia |
| Hypertension (systolic)^†^ (mm Hg) | 140– 159 | 160 – 179 | >155 | ER visit or hospitalization for malignant hypertension |
| Hypertension (diastolic)^†^(mm Hg) | 90 – 99 | 100– 109 | >100 | ER visit or hospitalization for malignant hypertension |
| Hypotension (systolic) (mm Hg) | 85 – 89 | 80 – 84 | <80 | ER visit or hospitalization for hypotensive shock |
| Respiratory rate (breaths per minute) | 17 – 20 | 21 – 25 | >25 | Intubation |
| Note: Subject should be at rest for all vital sign measurements.  ^*^ When resting heart rate is between 60 and 100 bpm. Use clinical judgement when characterizing bradycardia among some healthy subject populations (eg, conditioned athletes).  Source: DHHS 2007.^23^  ^†^Adapted from the South African Hypertension Society Hypertension Practice Guideline definitions.^24,25^ | | | | |

##### Table S5. Efficacy Endpoint Definitions of Covid-19 Severity.

| **COVID-19 Severity** | **Endpoint Definitions** |
| --- | --- |
| **Virologically Confirmed** | ≥1 COVID-19 disease symptom in Table S5  **AND**  Does not meet criteria for mild, moderate, or severe disease |
| **Mild** | ≥1 of:   - Fever (defined by subjective or objective measure, regardless of use of anti‑pyretic medications) - New onset cough - ≥2 Covid-19 respiratory/non-respiratory symptoms in Table S5   **AND**   - Does not meet criteria for moderate or severe |
| **Moderate** | ≥1 of:   - Fever (defined by subjective or objective measure, regardless of use of anti‑pyretic medications) + any 2 Covid-19 symptoms in Table S5 for ≥3 days (need not be contiguous days) - High fever (≥38.4°C) for ≥ 3 days (need not be contiguous days) - Any evidence of significant LRTI: - Shortness of breath (or breathlessness or difficulty breathing) with or without exertion (greater than baseline) - Tachypnea: 20 to 29 breaths per minute at rest - SpO_2_: 94% to 95% on room air - Abnormal chest x-ray or chest CT consistent with pneumonia or LRTI - Adventitious sounds on lung auscultation (eg, crackles/rales, wheeze, rhonchi, pleural rub, stridor)   **AND**   - Does not meet criteria for severe disease |
| **Severe** | ≥1 of:   - Tachypnea: ≥30 breaths per minute at rest - Resting heart rate ≥125 beats per minute - SpO_2_: ≤93% on room air or PAO_2_/FiO_2_ <300 - High flow oxygen therapy or NIV/NIPPV (eg, CPAP or BiPAP) - Mechanical ventilation or ECMO - One or more major organ system dysfunction or failure (eg, cardiac/circulatory, pulmonary, renal, hepatic, and/or neurological, to be defined by diagnostic testing/clinical syndrome/interventions), including any of the following:   - ARDS   - Acute renal failure   - Acute hepatic failure   - Acute right or left heart failure   - Septic or cardiogenic shock (with shock defined as SBP <90 mm Hg OR DBP <60 mm Hg   - Acute stroke (ischemic or hemorrhagic)   - Acute thrombotic event: AMI, DVT, PE   - Requirement for: vasopressors, systemic corticosteroids, or hemodialysis. - Admission to an ICU - Death |

Abbreviations: AMI = acute myocardial infarction; ARDS = acute respiratory distress syndrome; BiPAP = bi-level positive airway pressure; COVID-19 = coronavirus disease 2019; CPAP = continuous positive air pressure; CT = computed tomography; DBP = diastolic blood pressure; DVT = deep vein thrombosis; ECMO = extracorporeal membrane oxygenation; FiO_2_ = fraction of inspired oxygen; ICU = intensive care unit; LRTI = lower respiratory tract infection; NIV = non-invasive ventilation; NIPPV = non-invasive positive pressure ventilation; PAO_2_ = partial pressure of oxygen in the alveolus; PE = pulmonary embolism; SBP = systolic blood pressure; SpO_2_ = oxygen saturation.

##### Table S6. Symptoms of Suspected Covid-19.

| **Respiratory** | **Non-Respiratory** |
| --- | --- |
| New onset cough | Fever or feverishness (defined subjectively, or objective fever ≥37.8°C, regardless of use of anti‑pyretic medications) |
| New onset rapid breathing | Myalgia (or muscle ache) |
| New onset shortness of breath (or breathlessness or difficulty breathing) | Chills |
| Sore throat | Loss of taste (or taste disturbance) |
| Loss of smell (or smell disturbance) | Headache |
| Nasal congestion | Diarrhea |
| Runny nose | Tiredness (or fatigue or weakness) |
|  | Nausea or vomiting |
|  | Loss of appetite |

##### Table S7. Study Vaccine Groups With Maximized Immunization Plan.

| Cohorts/  Study Vaccine Groups | Number of  Randomized Subjects | Up to 2 Vaccinations | | |
| --- | --- | --- | --- | --- |
|  |  | Day 0 | | Day 21 |
|  |  | Stage 1^*^ | Stage 2^*^ | (+ 7 days) |
| **Cohort 1: HIV-negative subjects** | | | | |
| NVX-CoV2373 | N = 1480-2082^†^ | 444 | 1036-1638 | 1480-1638 |
| Placebo | N = 1480-2082^†^ | 444 | 1036-1638 | 1480-1638 |
| **Cohort 2: HIV-positive subjects** | | | | |
| NVX-CoV2373 | N = 120 | 40^‡^ | 80 | 120 |
| Placebo | N = 120 | 40^‡^ | 80 | 120 |

Abbreviations: HIV = human immunodeficiency virus; NVX-CoV2373 = 5 µg SARS-CoV-2 recombinant spike protein with 50 µg Matrix-M1 adjuvant; SMC = Safety Monitoring Committee; US = United States.

^*^	Vaccination of each cohort was divided into two stages for purposes of safety. Stage 1 of each cohort first vaccinated approximately 888 subjects aged 18 to <65 years. The SMC reviewed safety data through Day 7 in Stage 1 of Cohort 1 to determine progression to Stage 2 of Cohort 1, which vaccinated the remaining subjects aged 18 to <85 years in Cohort 1. NOTE: subjects aged ≥65 to <85 years were only enrolled during Stage 2 of Cohort 1.

^†^	A maximum of approximately 2082 subjects per vaccine group in Cohort 1 were to be enrolled (ie, up to a total of 4164 subjects in Cohort 1).

^‡^	Stage 1 enrollment for Cohort 2 began concurrently with Stage 2 enrollment of Cohort 1.

##### Figure S2. Active and Passive Surveillance Schedules.

**RESUME**

- Every 1-weekly ACTIVE SURVEILLANCE calls to FOLLOW-UP until resolution of symptoms; then resume every 2-weekly calls

**SCHEDULE**

- INITIAL Surveillance Visit **asap** within 1-3 days

**INSTRUCT**

- Subject to begin completing FLU-PRO questionnaire immediately for 10 days

**RESUME**

- Every 2-weekly ACTIVE SURVEILLANCE calls to ask about new onset of symptoms

**INITIAL** Report of symptoms

**No**

**Yes**

**ACTIVE SURVEILLANCE** (PHONE CALL w/ Script)

Recurring **OUTBOUND** calls from site to subject

- Every 2 weeks for new report of symptoms, **OR**
- Every 1 week for follow-up report of previous/ongoing symptoms

**PASSIVE SURVEILLANCE** (PHONE CALL w/ Script)

Spontaneous **INBOUND** calls from subject to site at any time

- New report of symptoms, **OR**
- Follow-up report of previous/ongoing symptoms

**FOLLOW-UP** Report of previous/ongoing symptoms

**ASK**

- DATE symptoms started?
- Which symptoms?
- Medical attention sought?
- New medications?
- Days of work missed?

**REMIND**

- Subject to call site at any time if experiencing new suspected COVID-19 symptoms

**ONGOING** suspect COVID-19 symptoms?

**ASK**

- Which symptoms still ongoing?
- Medical attention sought?
- New medications?
- Days of work missed?

**ASK**

- DATE symptoms resolved?
- Medical attention sought?
- New medications?
- Days of work missed?

**REMIND**

- Subject to call site at any time if visit to Emergency room / hospitalized

**REMIND**

- Subject to call site at any time if visit to Emergency room / hospitalized

**RESUME**

- Every 2-weekly ACTIVE SURVEILLANCE calls to ask about new onset of symptoms

**Yes**

**No**

**NEW** onset of suspected COVID-19

##### Figure S3. Initial and Follow-Up Surveillance Visits.

**RESUME**

- Every 1-weekly ACTIVE SURVEILLANCE calls to FOLLOW-UP until resolution of symptoms; then resume 2-weekly calls

**RESUME**

- Every 1-weekly ACTIVE SURVEILLANCE calls to FOLLOW-UP until resolution of symptoms; then resume 2-weekly calls

**INITIAL Surveillance VISIT (at STUDY site) – asap within 1-3 days**

- Record symptoms of suspected COVID-19 and onset date
- Vital signs (including MightySat Fingertip Pulse Oximeter)
- Lung exam (auscultation by qualified physician or nurse)
- New medications?
- Medical attention sought?
- Collect nasal swab for SARS-CoV-2 PCR testing

**PCR RESULT (SARS-CoV-2)**

**FOLLOW-UP Surveillance VISIT (HOME visit) – 4 days later (+ 2)**

- Record any ongoing symptoms of suspected COVID-19
- Vital signs (including MightySat Fingertip Pulse Oximeter)
- Lung exam (auscultation by qualified physician or nurse)
- New medications?
- Medical attention sought?
- DO NOT COLLECT a second nasal swab for SARS-CoV-2 PCR testing

**FOLLOW-UP Surveillance VISIT (at STUDY site) – 4 days later (+ 2)**

- Record any ongoing symptoms of suspected COVID-19
- Vital signs (including MightySat Fingertip Pulse Oximeter)
- Lung exam (auscultation by qualified physician or nurse)
- New medications?
- Medical attention sought?
- COLLECT REPEAT nasal swab for SARS-CoV-2 PCR testing

**NEW Onset of Symptoms** reported on **ACTIVE or PASSIVE Surveillance Phone Call**

**NOTE:**

- If subject is unable to visit clinic for INITIAL or FOLLOW-UP surveillance visit due to being hospitalized, then a qualified site staff (physician or nurse) will make every effort to visit subject in hospital to perform surveillance visit.
- In addition, following any emergency room and/or hospitalization visit, please request subject’s medical records and complete the study-specific hospitalization/ emergency room data collection form

**PCR positive**

**PCR negative**

### Supplemental Study Results:

#### Safety

Preliminary safety data were available on 968 participants at the time of the primary efficacy analysis, with safety presented by baseline serostatus (665 seronegative and 303 seropositive) to SARS‑CoV-2 infection.

##### Local Reactogenicity

Solicited local adverse events were predominantly mild to moderate and transient, and more common in vaccine recipients (Table S8). In baseline seronegative participants, injection site pain and tenderness were the most frequently reported solicited local adverse events following first (37% and 15% for pain and 25% and 11% for tenderness in vaccine and placebo recipients, respectively) and second (37% and 10% for pain and 25% and 8% for tenderness, respectively), with no increased frequency of events following second vaccination (Table S9). Similar results were seen in baseline seropositive participants. Severe local adverse events were infrequent but occurred more often in the vaccine group after second vaccination, but only in baseline seronegative participants.

##### Systemic Reactogenicity

Solicited systemic adverse events were predominantly mild to moderate and transient, and generally more common in vaccine recipients (Table S8). In baseline seronegative participants, headache, muscle pain, and fatigue were the most frequently reported solicited systemic adverse events following first (25% and 21% for headache in vaccine and placebo recipients, respectively; 18% and 10% for muscle pain; and 16% and 12% for fatigue) and second (20% and 19% for headache; 17% and 9% for muscle pain; and 13% and 7% for fatigue) vaccination, with no increased frequency of events following second vaccination (Table S9). Higher rates of systemic adverse events were generally reported in both vaccine and placebo recipients in baseline seropositive participants. Severe systemic adverse events were infrequent but generally occurred more often in the vaccine group after second vaccination. Fever was reported in few vaccine recipients.

##### Unsolicited Adverse Events

Medically attended adverse events and serious adverse events were infrequent but occurred slightly more often in the vaccine group, with no apparent clustering of specific adverse events by treatment group or system organ class (Table S10). To date, no serious adverse events have been assessed as related to trial vaccine by study investigators. No prespecified vaccination pause rules were triggered (Table S11).

##### Table S8. Summary of Adverse Events Among Stage 1 Trial Participants Through Day 35.

| **Parameters** | | **968 Stage 1 Participants (889 Healthy Adults and 79 PLWH)** | | | |
| --- | --- | --- | --- | --- | --- |
|  |  | **Baseline Seronegative** | | **Baseline Seropositive** | |
|  |  | **NVX-CoV2373** | **Placebo** | **NVX-CoV2373** | **Placebo** |
| **Participants in Group N1/N2^*^** | | **334/329** | **331/319** | **150/142** | **153/151** |
| **Solicited AEs^†^** | | | | | |
| Solicited local AEs | Dose 1 | 133 (39.8) | 63 (19.0) | 64 (42.7) | 33 (21.6) |
|  | Dose 2 | 129 (39.2) | 38 (11.9) | 56 (39.4) | 26 (17.2) |
| Severe local AEs | Dose 1 | 4 (1.2) | 2 (0.6) | 6 (4.0) | 1 (0.7) |
|  | Dose 2 | 14 (4.3) | 1 (0.3) | 5 (3.5) | 1 (0.7) |
| Solicited systemic AEs | Dose 1 | 130 (38.9) | 104 (31.4) | 58 (38.7) | 63 (41.2) |
|  | Dose 2 | 114 (34.7) | 89 (27.9) | 48 (33.8) | 42 (27.8) |
| Severe systemic AEs | Dose 1 | 7 (2.1) | 7 (2.1) | 7 (4.7) | 8 (5.2) |
|  | Dose 2 | 13 (4.0) | 14 (4.4) | 8 (5.6) | 7 (4.6) |
| **Unsolicited AEs^‡^** | | | | | |
| Any unsolicited AEs | Overall | 59 (17.7) | 54 (16.3) | 30 (20.0) | 38 (24.8) |
| Related unsolicited AEs | Overall | 10 (3.0) | 5 (1.5) | 3 (2.0) | 8 (5.2) |
| Severe unsolicited AEs | Overall | 1 (0.3) | 2 (0.6) | 3 (2.0) | 1 (0.7) |
| Severe/related unsolicited AEs | Overall | 0 | 0 | 0 | 0 |
| SAEs | Overall | 1 (0.3) | 1 (0.3) | 1 (0.7) | 0 |
| MAAEs | Overall | 9 (2.7) | 5 (1.5) | 4 (2.7) | 1 (0.7) |

Abbreviations: AE = adverse event; MAAE = medically attended adverse event; NVX-CoV2373 = 5 µg SARS-CoV-2 recombinant spike protein with 50 µg Matrix-M1 adjuvant; PLWH = people living with human immunodeficiency virus; SAE = serious adverse event.

^*^	N1/N2 represents total number of participants for Dose 1/Dose 2 for solicited AEs, with N1 representing the total number of participants for unsolicited AEs.

^†^	Solicited AEs were reported by participants (via diary or spontaneously) and had a recorded start date within the 7-day post‑vaccination window (ie, from trial Day 0 through Day 6).

^‡^	Safety follow-up from Day 35 through Day 385 is ongoing and remains blinded. To protect the integrity of safety data collection, data reported here have been provided to the sponsor’s clinical and regulatory personnel at the treatment group level only.

Note: Participants with multiple events within a category were counted only once, using the event with the greatest severity and/or relationship (Possible, Probably, Definite) as applicable.

##### Table S9. Summary of Local and Systemic Solicited Adverse Events From Day 0 Post-Vaccination to Day 6 (Post-Dose 1), and Day 21 Vaccination to Day 27 (Post-Dose 2), Inclusive – Safety Population.

| **Parameters** | | **968 Stage 1 Participants (889 Healthy Adults and 79 PLWH)** | | | |
| --- | --- | --- | --- | --- | --- |
|  |  | **Baseline Seronegative** | | **Baseline Seropositive** | |
|  |  | **NVX-CoV2373*** | **Placebo** | **NVX-CoV2373^*^** | **Placebo** |
| **Participants in Group N1/N2^†^** | | **334/329** | **331/319** | **150/142** | **153/151** |
| **Solicited Local AEs – Any Diary Day** | | | | | |
| All local AEs | Dose 1 | 133 (39.8) | 63 (19.0) | 64 (42.7) | 33 (21.6) |
|  | Dose 2 | 129 (39.2) | 38 (11.9) | 56 (39.4) | 26 (17.2) |
| *Grade 3* | Dose 1 | 4 ( 1.2) | 2 ( 0.6) | 6 ( 4.0) | 1 ( 0.7) |
|  | Dose 2 | 14 ( 4.3) | 1 ( 0.3) | 5 ( 3.5) | 1 ( 0.7) |
| Pain | Dose 1 | 123 (36.8) | 48 (14.5) | 58 (38.7) | 25 (16.3) |
|  | Dose 2 | 120 (36.5) | 31 ( 9.7) | 52 (36.6) | 22 (14.6) |
| *Grade 3* | Dose 1 | 3 ( 0.9) | 0 | 4 ( 2.7) | 1 ( 0.7) |
|  | Dose 2 | 11 ( 3.3) | 1 ( 0.3) | 3 ( 2.1) | 1 ( 0.7) |
| Tenderness | Dose 1 | 82 (24.6) | 37 (11.2) | 37 (24.7) | 23 (15.0) |
|  | Dose 2 | 83 (25.2) | 26 ( 8.2) | 29 (20.4) | 15 ( 9.9) |
| *Grade 3* | Dose 1 | 2 ( 0.6) | 1 ( 0.3) | 5 ( 3.3) | 0 |
|  | Dose 2 | 10 ( 3.0) | 0 | 3 ( 2.1) | 0 |
| Erythema | Dose 1 | 5 ( 1.5) | 2 ( 0.6) | 2 ( 1.3) | 0 |
|  | Dose 2 | 6 ( 1.8) | 0 | 0 | 0 |
| *Grade 3* | Dose 1 | 0 | 1 ( 0.3) | 0 | 0 |
|  | Dose 2 | 1 ( 0.3) | 0 | 0 | 0 |
| Swelling | Dose 1 | 4 ( 1.2) | 2 ( 0.6) | 6 ( 4.0) | 0 |
|  | Dose 2 | 9 ( 2.7) | 0 | 1 ( 0.7) | 2 ( 1.3) |
| *Grade 3* | Dose 1 | 0 | 1 ( 0.3) | 0 | 0 |
|  | Dose 2 | 1 ( 0.3) | 0 | 0 | 0 |
| **General Systemic AEs – Any Diary Day** | | | | | |
| All systemic AEs | Dose 1 | 130 (38.9) | 104 (31.4) | 58 (38.7) | 63 (41.2) |
|  | Dose 2 | 114 (34.7) | 89 (27.9) | 48 (33.8) | 42 (27.8) |
| *Grade 3* | Dose 1 | 7 ( 2.1) | 7 ( 2.1) | 7 ( 4.7) | 8 ( 5.2) |
|  | Dose 2 | 13 ( 4.0) | 14 ( 4.4) | 8 ( 5.6) | 7 ( 4.6) |
| Temperature | Dose 1 | 4 ( 1.2)^‡^ | 4 ( 1.2) | 5 ( 3.4)^§^ | 3 ( 2.0) |
|  | Dose 2 | 8 ( 2.4) | 2 ( 0.6) | 9 ( 6.3) | 5 ( 3.3) |
| *Grade 3* | Dose 1 | 1 ( 0.3)^‡^ | 1 ( 0.3) | 0^§^ | 1 ( 0.7) |
|  | Dose 2 | 0 | 0 | 1 ( 0.7) | 1 ( 0.7) |
| Headache | Dose 1 | 83 (24.9) | 70 (21.1) | 35 (23.3) | 44 (28.8) |
|  | Dose 2 | 67 (20.4) | 59 (18.5) | 30 (21.1) | 30 (19.9) |
| *Grade 3* | Dose 1 | 3 ( 0.9) | 1 ( 0.3) | 2 ( 1.3) | 7 ( 4.6) |
|  | Dose 2 | 6 ( 1.8) | 8 ( 2.5) | 5 ( 3.5) | 5 ( 3.3) |
| Fatigue | Dose 1 | 52 (15.6) | 38 (11.5) | 23 (15.3) | 21 (13.7) |
|  | Dose 2 | 51 (15.5) | 35 (11.0) | 17 (12.0) | 18 (11.9) |
| *Grade 3* | Dose 1 | 0 | 3 ( 0.9) | 5 ( 3.3) | 2 ( 1.3) |
|  | Dose 2 | 4 ( 1.2) | 4 ( 1.3) | 1 ( 0.7) | 3 ( 2.0) |
| Malaise | Dose 1 | 31 ( 9.3) | 27 ( 8.2) | 21 (14.0) | 17 (11.1) |
|  | Dose 2 | 28 ( 8.5) | 24 ( 7.5) | 18 (12.7) | 14 ( 9.3) |
| *Grade 3* | Dose 1 | 1 ( 0.3) | 3 ( 0.9) | 3 ( 2.0) | 2 ( 1.3) |
|  | Dose 2 | 2 ( 0.6) | 2 ( 0.6) | 2 ( 1.4) | 2 ( 1.3) |
| Joint pain | Dose 1 | 41 (12.3) | 22 ( 6.6) | 26 (17.3) | 19 (12.4) |
|  | Dose 2 | 43 (13.1) | 21 ( 6.6) | 22 (15.5) | 13 ( 8.6) |
| *Grade 3* | Dose 1 | 3 ( 0.9) | 1 ( 0.3) | 2 ( 1.3) | 1 ( 0.7) |
|  | Dose 2 | 4 ( 1.2) | 0 | 3 ( 2.1) | 1 ( 0.7) |
| Muscle pain | Dose 1 | 59 (17.7) | 32 ( 9.7) | 30 (20.0) | 17 (11.1) |
|  | Dose 2 | 56 (17.0) | 27 ( 8.5) | 26 (18.3) | 11 ( 7.3) |
| *Grade 3* | Dose 1 | 4 ( 1.2) | 2 ( 0.6) | 4 ( 2.7) | 2 ( 1.3) |
|  | Dose 2 | 5 ( 1.5) | 4 ( 1.3) | 4 ( 2.8) | 1 ( 0.7) |
| **Gastrointestinal Systemic AEs** | | | | | |
| Nausea or vomiting | Dose 1 | 27 ( 8.1) | 17 ( 5.1) | 14 ( 9.3) | 14 ( 9.2) |
|  | Dose 2 | 19 ( 5.8) | 20 ( 6.3) | 13 ( 9.2) | 16 (10.6) |
| *Grade 3* | Dose 1 | 0 | 0 | 2 ( 1.3) | 1 ( 0.7) |
|  | Dose 2 | 0 | 1 ( 0.3) | 1 ( 0.7) | 2 ( 1.3) |

Abbreviations: AE = adverse event; CI = confidence interval; FDA = US Food and Drug Administration; HIV = human immunodeficiency virus; NVX-CoV2373 = 5 µg SARS-CoV-2 recombinant spike protein with 50 µg Matrix-M1 adjuvant; PLWH = people living with human immunodeficiency virus.

^*^ Includes Matrix-M1 adjuvant.

^†^	N1/N2 represents total number of participants for Dose 1/Dose 2.

^‡^	N1=333.

^§^N1=148.

Note: At each level of subject summarization, a participant was counted once if the subject reported one or more events. The 95% CI was based on the exact Clopper-Pearson method. Toxicity grading was standardized according to the FDA toxicity grading scale.

Note: Values are represented as n (%).

##### Table S10. Summary of Unsolicited Treatment-Emergent Medically Attended Adverse Events Through 35 Days After First Vaccination (Safety Analysis Set).

| **System Organ Class/Preferred Term^*^** | **968 Stage 1 Participants (889 Healthy Adults and 79 PLWH)** | | | |
| --- | --- | --- | --- | --- |
|  | **Baseline Seronegative** | | **Baseline Seropositive** | |
|  | **NVX-CoV2373** | **Placebo** | **NVX-CoV2373** | **Placebo** |
| **Participants in Group N** | **334** | **331** | **150** | **153** |
| Any MAAE | 9 ( 2.7) | 5 ( 1.5) | 4 ( 2.7) | 1 ( 0.7) |
| Infections and infestations | 3 ( 0.9) | 3 ( 0.9) | 2 ( 1.3) | 0 |
| Abscess limb | 0 | 0 | 2 ( 1.3) | 0 |
| Conjunctivitis | 0 | 1 ( 0.3) | 0 | 0 |
| Infected bite | 1 ( 0.3) | 0 | 0 | 0 |
| Lung abscess | 0 | 1 ( 0.3) | 0 | 0 |
| Tonsillitis | 1 ( 0.3) | 0 | 0 | 0 |
| Upper respiratory tract infection | 1 ( 0.3) | 0 | 0 | 0 |
| Urinary tract infection | 0 | 1 ( 0.3) | 0 | 0 |
| Injury, poisoning and procedural complications | 1 ( 0.3) | 1 ( 0.3) | 2 ( 1.3) | 0 |
| Contusion | 1 ( 0.3) | 0 | 0 | 0 |
| Facial bones fracture | 0 | 0 | 1 ( 0.7) | 0 |
| Skin laceration | 0 | 0 | 1 ( 0.7) | 0 |
| Thermal burn | 0 | 1 ( 0.3) | 0 | 0 |
| Skin and subcutaneous tissue disorders | 2 ( 0.6) | 0 | 0 | 1 ( 0.7) |
| Dermatitis | 1 ( 0.3) | 0 | 0 | 0 |
| Papule | 0 | 0 | 0 | 1 ( 0.7) |
| Pityriasis rosea | 1 ( 0.3) | 0 | 0 | 0 |
| Blood and lymphatic system disorders | 1 ( 0.3) | 0 | 0 | 0 |
| Anemia | 1 ( 0.3) | 0 | 0 | 0 |
| Gastrointestinal disorders | 1 ( 0.3) | 0 | 0 | 0 |
| Abdominal pain | 1 ( 0.3) | 0 | 0 | 0 |
| General disorders and administration site conditions | 0 | 1 ( 0.3) | 0 | 0 |
| Influenza like illness | 0 | 1 ( 0.3) | 0 | 0 |
| Musculoskeletal and connective tissue disorders | 0 | 1 ( 0.3) | 0 | 0 |
| Arthralgia | 0 | 1 ( 0.3) | 0 | 0 |
| Nervous system disorders | 0 | 1 ( 0.3) | 0 | 0 |
| Headache | 0 | 1 ( 0.3) | 0 | 0 |
| Renal and urinary disorders | 1 ( 0.3) | 0 | 0 | 0 |
| Urethral discharge | 1 ( 0.3) | 0 | 0 | 0 |
| Respiratory, thoracic and mediastinal disorders | 0 | 1 ( 0.3) | 0 | 0 |
| Nasal congestion | 0 | 1 ( 0.3) | 0 | 0 |
| Oropharyngeal pain | 0 | 1 ( 0.3) | 0 | 0 |

Abbreviations: MAAE = medically attended adverse event; MedDRA = *Medical Dictionary for Regulatory Activities*; NVX‑CoV2373 = 5 µg SARS-CoV-2 recombinant spike protein with 50 µg Matrix-M1 adjuvant; PLWH = people living with human immunodeficiency virus.

^*^ Coded using MedDRA, Version 23.0.

##### Table S11. Summary of Unsolicited Serious Treatment-Emergent Adverse Events Through 35 Days After First Vaccination (Safety Analysis Set).

| **System Organ Class/Preferred Term^*^** | **968 Stage 1 Participants (889 Healthy Adults and 79 PLWH)** | | | |
| --- | --- | --- | --- | --- |
|  | **Baseline Seronegative** | | **Baseline Seropositive** | |
|  | **NVX-CoV2373** | **Placebo** | **NVX-CoV2373** | **Placebo** |
| **Participants in Group N** | **334** | **331** | **150** | **153** |
| Any SAE | 1 (0.3) | 1 (0.3) | 1 (0.7) | 0 |
| Blood and lymphatic system disorders | 1 (0.3) | 0 | 0 | 0 |
| Anemia | 1 (0.3) | 0 | 0 | 0 |
| Infections and infestations | 0 | 1 (0.3) | 0 | 0 |
| Lung abscess | 0 | 1 (0.3) | 0 | 0 |
| Injury, poisoning and procedural complications | 0 | 0 | 1 (0.7) | 0 |
| Facial bones fracture | 0 | 0 | 1 (0.7) | 0 |

Abbreviations: MedDRA = *Medical Dictionary for Regulatory Activities*; NVX‑CoV2373 = 5 µg SARS-CoV-2 recombinant spike protein with 50 µg Matrix-M1 adjuvant; PLWH = people living with human immunodeficiency virus; SAE = serious adverse event.

^*^ Coded using MedDRA, Version 23.0.

22.	Division of AIDS (DAIDS) NIoAaID, National Institutes of Health. Division of AIDS (DAIDS) table for grading the severity of adult and pediatric adverse events. In: Services UDoHaH, ed.July 2017.

23.	Food and Drug Administration CfBEaRU. Guidance for industry: Toxicity grading scale for healthy adult and adolescent volunteers enrolled in preventive vaccine clinical trials. In: (DHHS) DoHaHS, ed.September 2007.

24.	Hypertension guideline working g, Seedat YK, Rayner BL, Veriava Y. South African hypertension practice guideline 2014. Cardiovasc J Afr 2014;25:288-94.

25.	Rayner B, Jones E, Veriava Y, Seedat YK. South African Hypertension Society commentary on the American College of Cardiology/American Heart Association hypertension guidelines. Cardiovasc J Afr 2019;30:184-7.
